# Personalized Mood Prediction from Patterns of Behavior Collected with Smartphones

**DOI:** 10.1101/2022.10.12.22281007

**Authors:** Brunilda Balliu, Chris Douglas, Darsol Seok, Liat Shenhav, Yue Wu, Doxa Chatzopoulou, William Kaiser, Victor Chen, Jennifer Kim, Sandeep Deverasetty, Inna Arnaudova, Robert Gibbons, Eliza Congdon, Michelle G. Craske, Nelson Freimer, Eran Halperin, Sriram Sankararaman, Jonathan Flint

## Abstract

Over the last ten years, there has been considerable progress in using digital behavioral phenotypes, captured passively and continuously from smartphones and wearable devices, to infer depressive mood. However, most digital phenotype studies suffer from poor replicability, often fail to detect clinically relevant events, and use measures of depression that are not validated or suitable for collecting large and longitudinal data. Here, we report high-quality longitudinal validated assessments of depressive mood from computerized adaptive testing paired with continuous digital assessments of behavior from smartphone sensors for up to 40 weeks on 183 individuals experiencing mild to severe symptoms of depression. We apply a combination of cubic spline interpolation and idiographic models to generate individualized predictions of future mood from the digital behavioral phenotypes, achieving high prediction accuracy of depression severity up to three weeks in advance (R^2^ :2 80%) and a 65.7% reduction in the prediction error over a baseline model which predicts future mood based on past depression severity alone. Finally, our study verified the feasibility of obtaining high-quality longitudinal assessments of mood from a clinical population and predicting symptom severity weeks in advance using passively collected digital behavioral data. Our results indicate the possibility of expanding the repertoire of patient-specific behavioral measures to enable future psychiatric research.

## Introduction

Major depressive disorder (MDD) affects almost one in five people^1^ and is now the world’s leading cause of disability^2^. However, it is often undiagnosed: only about half of those with MDD are identified and offered treatment^3,4^. In addition, for many people, MDD is a chronic condition characterized by periods of relapse and recovery that requires ongoing monitoring of symptoms. MDD diagnosis and symptom monitoring is typically dependent on clinical interview, a method that rarely exceeds an inter-rater reliability of 0.7^5,6^. Furthermore, sufferers are unlikely to volunteer that they are depressed because of the reduced social contact associated with low mood and because of the stigma attached to admitting to being depressed. Developing new ways to quickly and accurately diagnose MDD or monitor depressive symptoms in real time would substantially alleviate the burden of this common and debilitating condition.

The advent of electronic methods of collecting information, e.g., smartphone sensors or wearable devices, means that behavioral measures can now be obtained as individuals go about their daily lives. Over the last ten years there has been considerable progress in using these digital behavioral phenotypes to infer mood and depression^7–15^. Yet, most digital mental health studies suffer from one or more of the following limitations^16–18^. First, many studies are likely underpowered to meet their analytic objectives^10,12,19,20^. Second, most studies do not follow up subjects long enough to adequately capture changes in signal within an individual over time^10,11,19,21,22^, even though such changes are highly informative for clinical care. The few studies with longitudinal assessments use ecological momentary assessments^19,20,23^ to measure state mood, rather than a psychometrically validated symptom scale for depression. Furthermore, they examine associations between behavior and mood at a population level^23^. This nomothetic approach is limited by the fact that both mood and its relationship to behavior can vary substantially between individuals. Last, many of the existing studies focus on healthy subjects, thus prohibiting evaluation of how well digital phenotypes perform in predicting depression^24^.

Here, we overcome these limitations by using a validated measure of depression from computerized adaptive testing^25^ to obtain high-quality longitudinal measures of mood. Computerized adaptive testing is a technology for interactive administration of tests that tailors the test to the examinee (or, in our application, to the patient)^26^. Tests are ‘adaptive’ in the sense that the testing is driven by an algorithm that selects questions in real time and in response to the on-going responses of the patient. By employing item response theory to select a small number of questions from a large bank, the test provides a powerful and efficient way to detect psychiatric illness without suffering response fatigue. We also use smartphone sensing^27^ to passively and continuously collect behavioral phenotypes for up to 40 weeks on 183 individuals experiencing mild to severe symptoms of depression (3,005 days with mood assessment and 29,254 days with behavioral assessment). To account for inter-individual heterogeneity and provide individual-specific predictors of depression trajectories we use an idiographic (or, personalized) modeling approach. Ultimately, we expect that this approach will provide patient-specific predictors of depressive symptom severity to guide personalized intervention, as well as enable future psychiatric research, for example in genome and phenome-wide association studies.

## Results

### Study participants and treatment protocol

Participants (N = 437; 76.5% female, 26.5% white) are University of California Los Angeles (UCLA) students experiencing mild to severe symptoms of depression or anxiety enrolled as part of the Screening and Treatment for Anxiety and Depression^28^ (STAND) study (Sup Figure 1). The STAND eligibility criteria and treatment protocol are described extensively elsewhere^29^. Briefly, participants are initially assessed using the Computerized Adaptive Testing Depression Inventory^31^ (CAT-DI), an online adaptive tool that offers validated assessments of depression severity (measured on a 0-100 scale). After the initial assessment, participants are routed to appropriate treatment resources depending on depression severity: those with mild (35 ≤ CAT-DI < 65) to moderate (65 ≤ CAT-DI < 75) depression at baseline received online support with or without peer coaching^30^ while those with severe depression (CAT-DI ≥ 75) received in-person care from a clinician (Materials and Methods).

**Figure 1:**
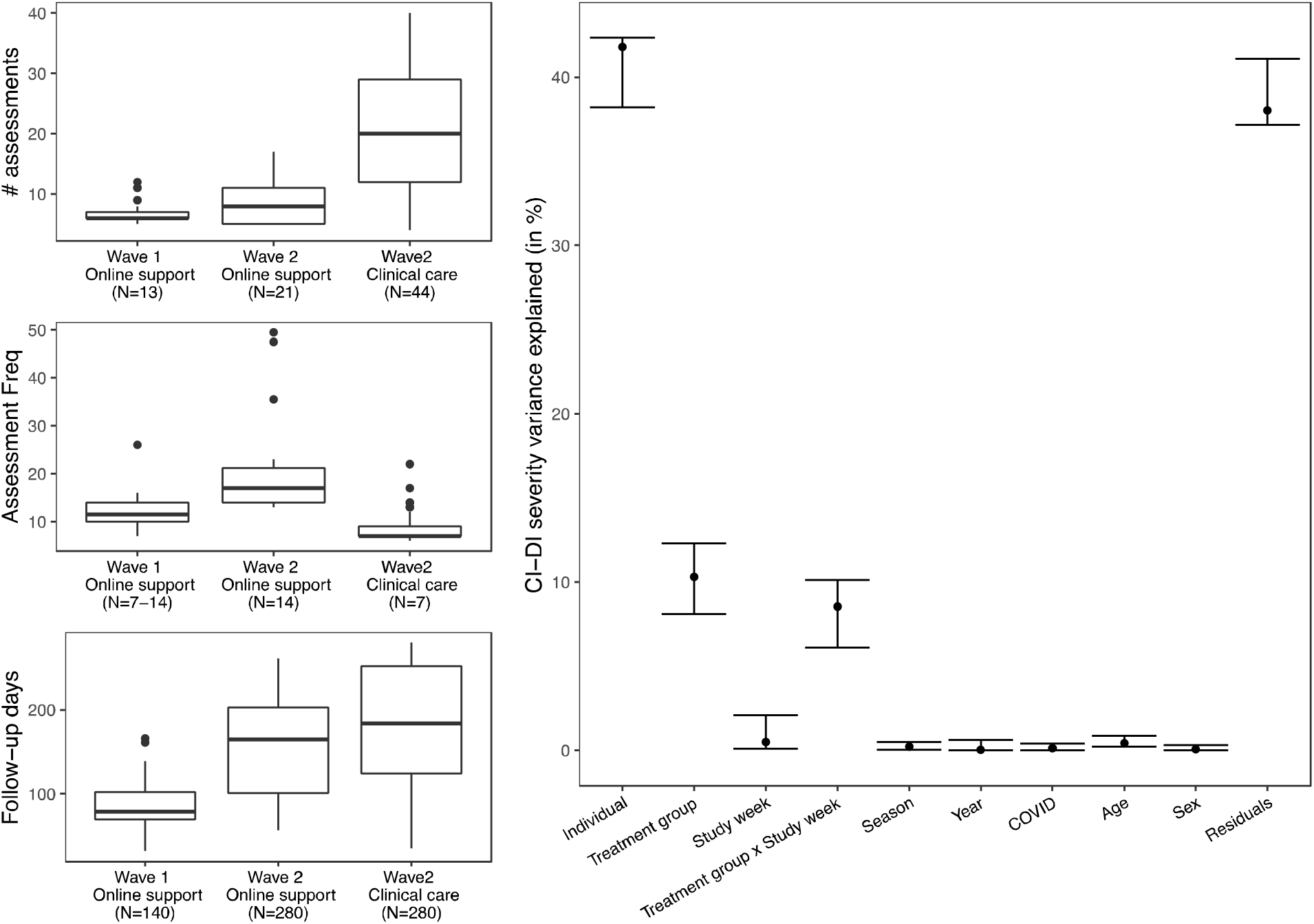
Overview of CAT-DI assessment frequency and source of variation in CAT-DI. **(a-c)** Box plot of the observed number of CAT-DI assessments (a), median number of days between assessments (b), and follow-up time in days (c) for each wave and treatment group. The numbers in the parentheses indicate the expected values for each of these metrics according to study design (Sup Figure 2). The dark black line represents the median value; the box limits show the interquartile range (IQR) from the first (Q1) to third (Q3) quartiles; the whiskers extend to the furthest data point within Q1-1.5*IQR (bottom) and Q3+1.5*IQR (top). **(d)** Proportion of CAT-DI severity variance explained (VE) by inter-individual differences and other study parameters with 95% confidence intervals. The proportion of variance attributable to each source was computed using a linear mixed model with the individual id and season (two multilevel categorical variables) modeled as random variables and all other variables modeled as fixed (see Materials and Methods).

STAND enrolled participants in two waves, each with different inclusion criteria and CAT-DI assessment and treatment protocol (Sup Figure 2a). Wave 1 was limited to individuals with mild to moderate symptoms at baseline (N=182) and treatment lasted for up to 20 weeks. Wave 2 included individuals with mild to moderate (N=142) and severe (N=124) symptoms and treatment lasted for up to 40 weeks. Eleven individuals participated in both waves. Depression symptom severity was assessed up to every other week for the participants that received online support (both waves), i.e., those with mild to moderate symptoms, and every week for the participants that received in-person clinical care, i.e., those with severe symptoms (Materials and Methods).

**Figure 2:**
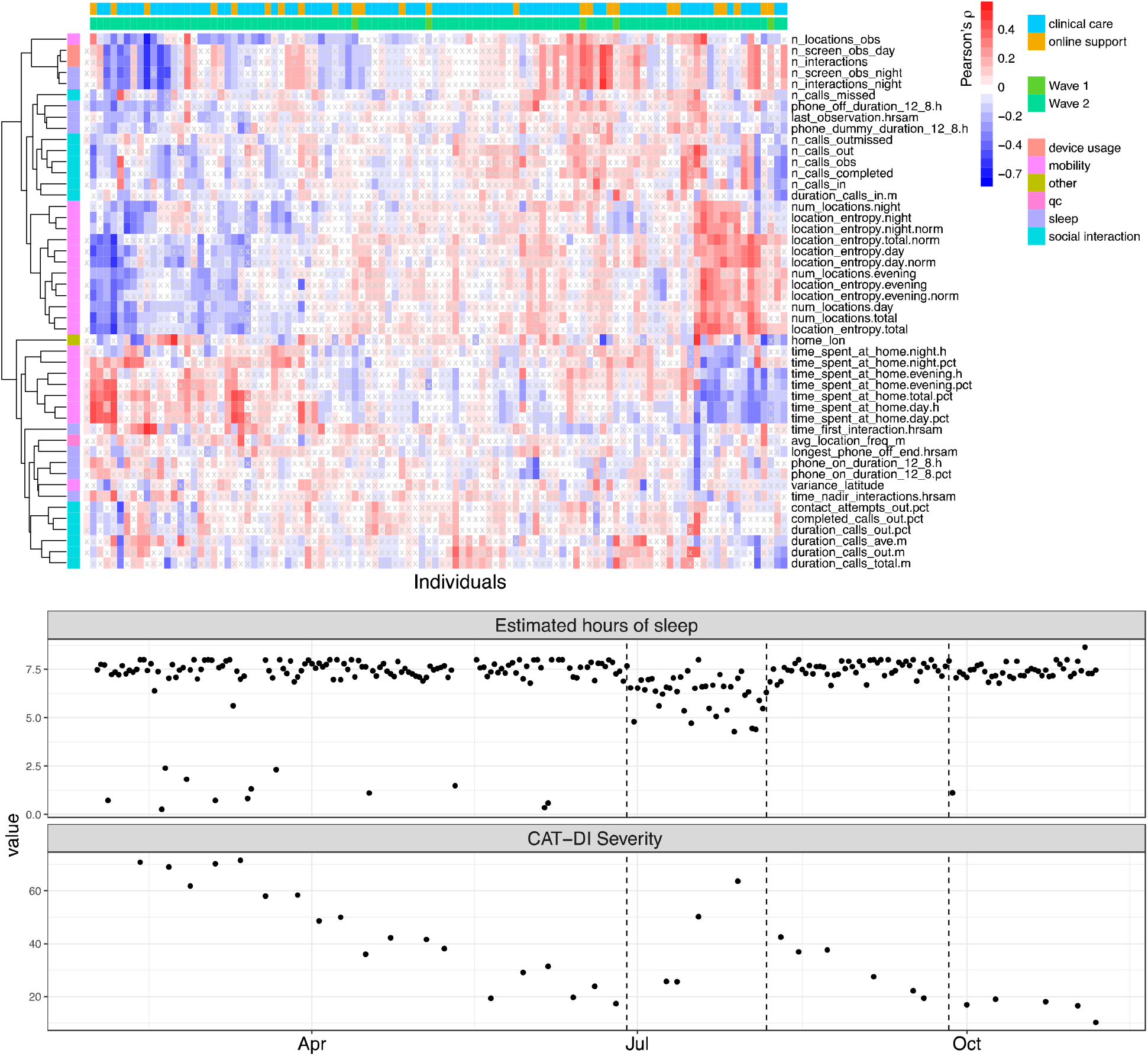
Overview of correlation between depression severity scores and features. **(a)** Heatmap for Pearson’s correlation coefficient (color of cell) between CAT-DI scores and behavioral features (y-axis) across individuals (first column) and within each individual (x-axis). Correlation coefficients with BH-adjusted p-values > 0.05 are indicated by x. For plotting ease, we limit to untransformed features (N=50, see Materials and Methods). Rows and columns are annotated by feature type and by each individual’s wave and treatment group. Rows and columns are ordered using hierarchical clustering with Euclidean distance. **(b)** Example of identifying window of potential sleep disruption using sensor data related to phone usage and screen on/off status. The top panel shows estimated hours of sleep for an individual during the study while the bottom panel shows the depression severity scores during the same period. The dotted lines indicate the dates at which a change point is estimated to have occurred in the estimated hours of sleep as estimated using a change point model framework for sequential change detection (Materials and Methods). BH: Benjamini Hochberg.

### Adherence to CAT-DI assessment protocol

Overall, participants provided a total of 4,507 CAT-DI assessments (out of 11,218 expected by the study protocols). Participant adherence to CAT-DI assessments varied across enrollment waves (Likelihood ratio test [LRT] P-value < 2.2×10^−16^), treatment groups (LRT P-value < 2.2×10^−16^), and during the follow-up period (LRT P-value = 1.29×10^−6^). Specifically, participants that received clinical care were more adherent than those which only received online support (Sup Figure 2b). Attrition for participants which received clinical care was linear over the follow-up period, with 1.7% of participants dropping out CAT-DI assessments within two weeks into the study. Attrition for participants that received online support was large two weeks into the study (33.5% of Wave 1 and 37.3% of Wave 2 participants) and linear for the remaining of the study.

Participant adherence to CAT-DI assessments varied with sex and age. Among participants that received online support, men were less likely to complete all CAT-DI assessments in wave 1 (OR= 0.86, LRT P-value = 2.9×10^−4^) but more likely to complete them in wave 2 (OR= 1.31, LRT P-value = 3.1×10^−11^). Participant adherence did not vary with sex for those receiving clinical support. In addition, among participants that received online support in wave 2, older participants were more likely to complete all CAT-DI assessments than younger participants (OR=1.13, LRT P-value < 2.2×10^−16^). Participant adherence did not vary with age for participants in wave 1 or those receiving clinical support in wave 2.

For building personalized mood prediction models, we focus on 183 individuals (49 from Wave 1 and 134 from Wave 2) who had at least five mental health assessments during the study (Materials and Methods). For these individuals we obtained a total of 3,005 CAT-DI assessments with a median of 13 assessments, 171 follow-up days, and 10 days between assessments per individual (Figure 1a-c).

### Computerized adaptive testing captures treatment-related changes in depression severity

We assessed what factors contribute to variation in the CAT-DI severity scores (Figure 1e, Materials and Methods). Subjects are assigned to different treatments (online support or clinical care) depending on their CAT-DI severity scores, so not surprisingly we see a significant source of variation attributable to the treatment group (10.3% of variance explained, 95% CI: 8.37 - 12.68%). Once assigned to a treatment group, we expect to see changes over time as treatment is delivered to individuals with severe symptoms at baseline. This is reflected in a significant source of variation attributable to the interaction between the treatment group and the number of weeks spent in the study (8.54% of variance explained, 95% CI: 5.92 - 10.4%) and the improved scores for individuals with severe symptoms at baseline as they spend more time in the study (Sup Figure 3). We found no statistically significant effect of the COVID pandemic, sex, and other study parameters. The largest source of variation in depression severity scores is attributable to between-individual differences (41.78% of variance explained, 95% CI: 38.31 - 42.02%), suggesting that accurate prediction of CAT-DI severity requires learning models tailored to each individual.

**Figure 3:**
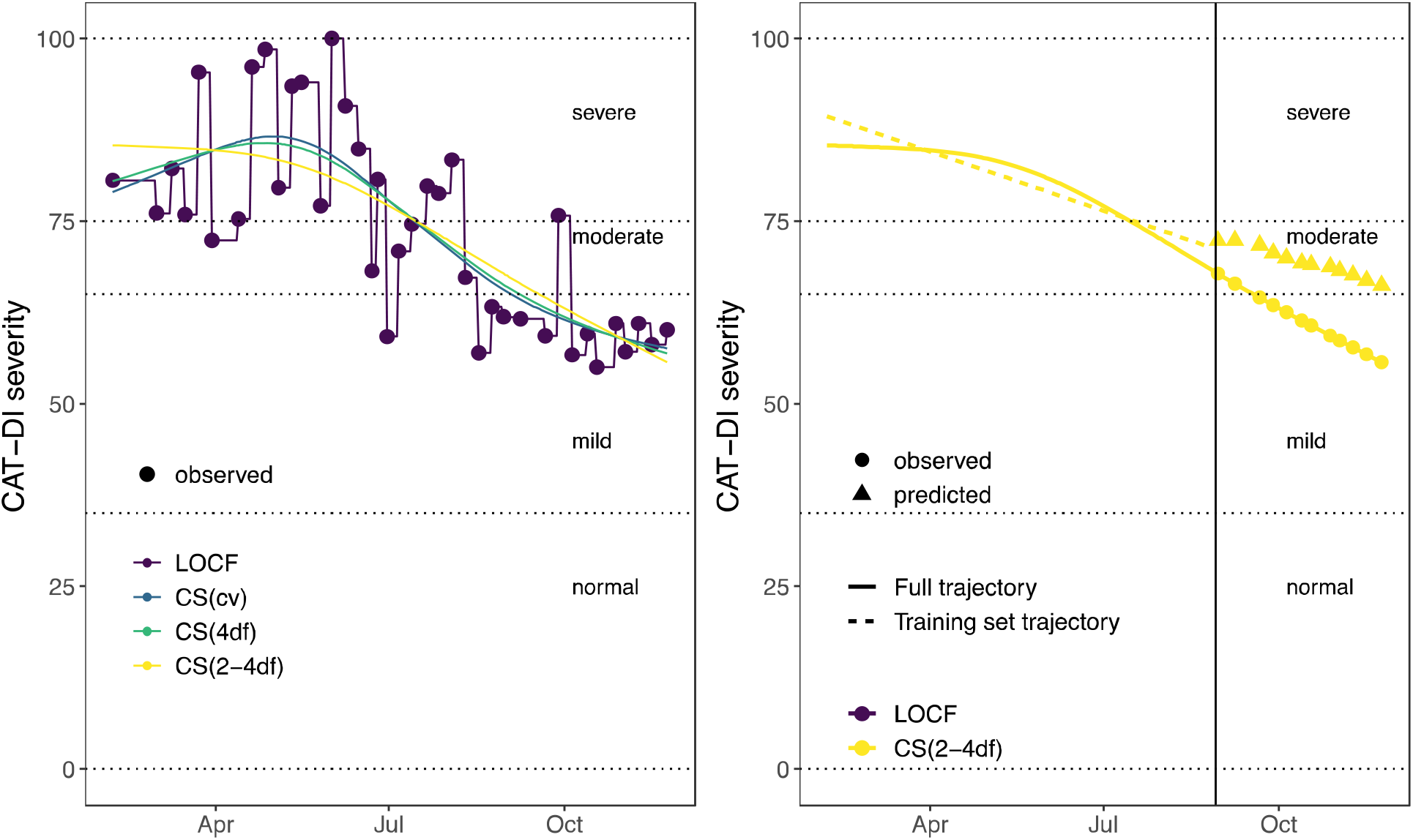
Interpolation of depression severity scores and latent trait inference. **(a)** Illustration of different interpolation methods considered for imputing the depression severity scores and inferring the latent depression traits. The dotted horizontal lines indicate the depression severity score thresholds for the normal (0 _≤_ CAT-DI < 35), mild (35 _≤_ CAT-DI < 65), moderate (65≤ CAT-DI < 75), and severe (75 _≤_ CAT-DI _≤_100) depression severity categories. **(b)** Illustration of the prediction method for the CS(2-4df) interpolation method. We first infer the latent trait on the full CAT-DI trajectory of an individual (continuous yellow line). We then split the trajectory into a training set (days 1 until t) and a test set (days t+1 until T), infer the latent trait on the training set (dashed yellow line), and predict the trajectory in the test set (yellow triangles). Finally, we compute prediction accuracy metrics by comparing the observed (yellow circles) and predicted (yellow triangles) depression scores in the test set. We follow a similar approach for the other interpolation methods. The vertical line indicates the first date of the test set trajectory, i.e., the last 30% of the trajectory. LOCF: last observation carried forward. CS (xdf): cubic spline with x degrees of freedom. CS (cv): best-fitting cubic spline according to leave-one-out cross-validation.

### Digital behavioral phenotypes capture changes in behavior

We set out to examine how digital behavioral phenotypes change over time for each person and with CAT-DI severity scores. For example, we want to know how hours of sleep on a specific day for a specific individual differs from the average hours of sleep in the previous week, or month. To answer these questions, we extracted digital behavioral phenotypes (referred to hereinafter as features) captured from participants’ smartphone sensors and investigated which features predicted the CAT-DI scores. STAND participants had the AWARE framework^27^ installed on their smartphones, which queried phone sensors to obtain information about a participant’s location, screen on/off behavior, and number of incoming and outgoing text messages and phone calls. We processed these measurements (Materials and Methods and Supplementary Material) to obtain daily aggregate measures of activity (23 features), social interaction (18 features), sleep quality (13 features), and device usage (two features). In addition, we processed these features to capture relative changes in each measure for each individual, e.g., changes in average amount of sleep in the last week compared to what is typical over the last month. In total, we obtained 1,325 features. Missing daily feature values (Sup Figure 4) were imputed using two different imputation methods, AutoComplete^31^ and softImpute^32^ (Materials and Methods), resulting in 29,254 days of logging events across all individuals.

**Figure 4:**
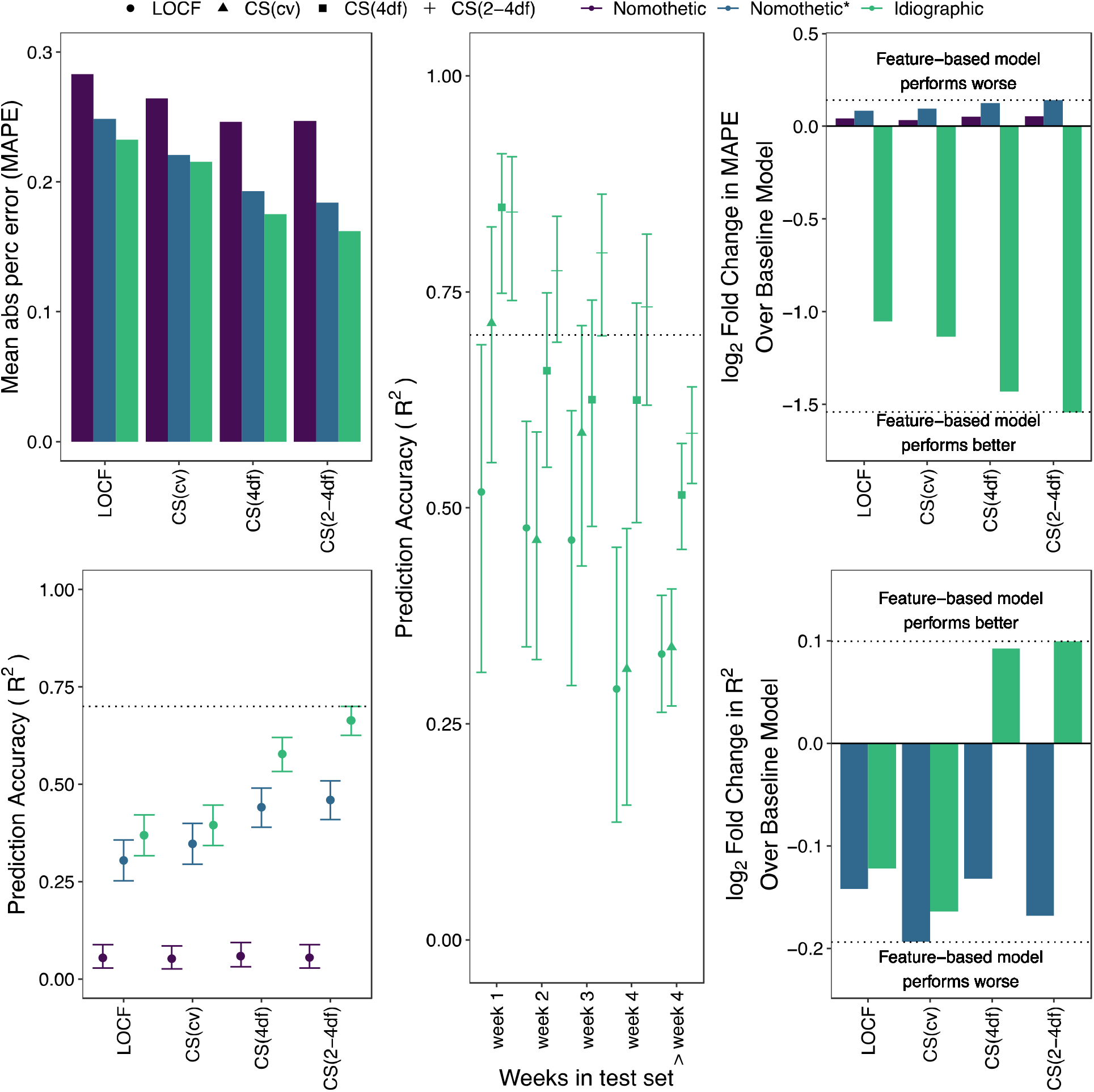
Idiographic models achieve higher group level prediction accuracy than nomothetic models. **(a-b)** CAT-DI prediction accuracy across all individuals in the test set as measured by MAPE (a) and R^2^ (b) across all individuals for different models and latent depression traits. The dotted line in B indicates 70% prediction accuracy and bars indicate 95% confidence intervals of R^2^. **(c)** R^2^ versus the number of weeks ahead we are predicting from the last observation in the training set. The dotted line indicates 70% prediction accuracy. Bars indicate 95% confidence intervals of R^2^. **(d-e)** log2 fold change in CAT-DI prediction accuracy, as measured by MAPE (d) and R^2^ (e), of feature-based model over the baseline model. Negative log2 fold change in MAPE and positive log2 fold change in R^2^ mean that the feature-based model performs better than the baseline model. A log2 fold change in MAPE of −1 means that the prediction error of the baseline model is twice as large as that of the feature-based model. The dotted line indicates the log2 fold change for the best and worst performing model/latent trait combination. Features were imputed with Autocomplete and CAT-DI was modeled using a logistic elastic net regression. MAPE: mean absolute percent error. LOCF: last observation carried forward. CS(xdf): cubic spline with x degrees of freedom. CS(cv): best-fitting cubic spline according to leave-one-out cross-validation.

Several of these features map onto the DSM-5 MDD criteria of anhedonia, sleep disturbance, and loss of energy (Supplementary Material; Sup Figure 5). We computed correlations between these features and an individuals’ depression severity score and found that these features often correlate strongly with changes in depression (Figure 2a). For example, for one individual, the number of unique locations visited during the day shows a strong negative correlation with their depression severity scores during the study (Pearson’s ρ = −0.65, Benjamini-Hochberg [BH]-adjusted P-value = 2.50×10^−20^). We observed a lot of heterogeneity in the strength and direction of the correlation of these features with depression severity across individuals. For example, features related to location entropy are positively (Pearson’s ρ=0.40, BH-adjusted P-value = 3.55×10^−11^) correlated with depression severity for some individuals but negatively (ρ=-0.59, BH-adjusted P-value = 1.11×10^−22^) or not correlated (ρ=-3.92×10^−04^, BH-adjusted P-value = 0.995) for others. Finally, as expected from the large heterogeneity in these correlation between individuals, the correlation of these features with depression severity scores across individuals was very poor, the strongest correlation was observed for the wake-up time (ρ=.07, BH-adjusted P-value = 2.47×10^−03^).

**Figure 5:**
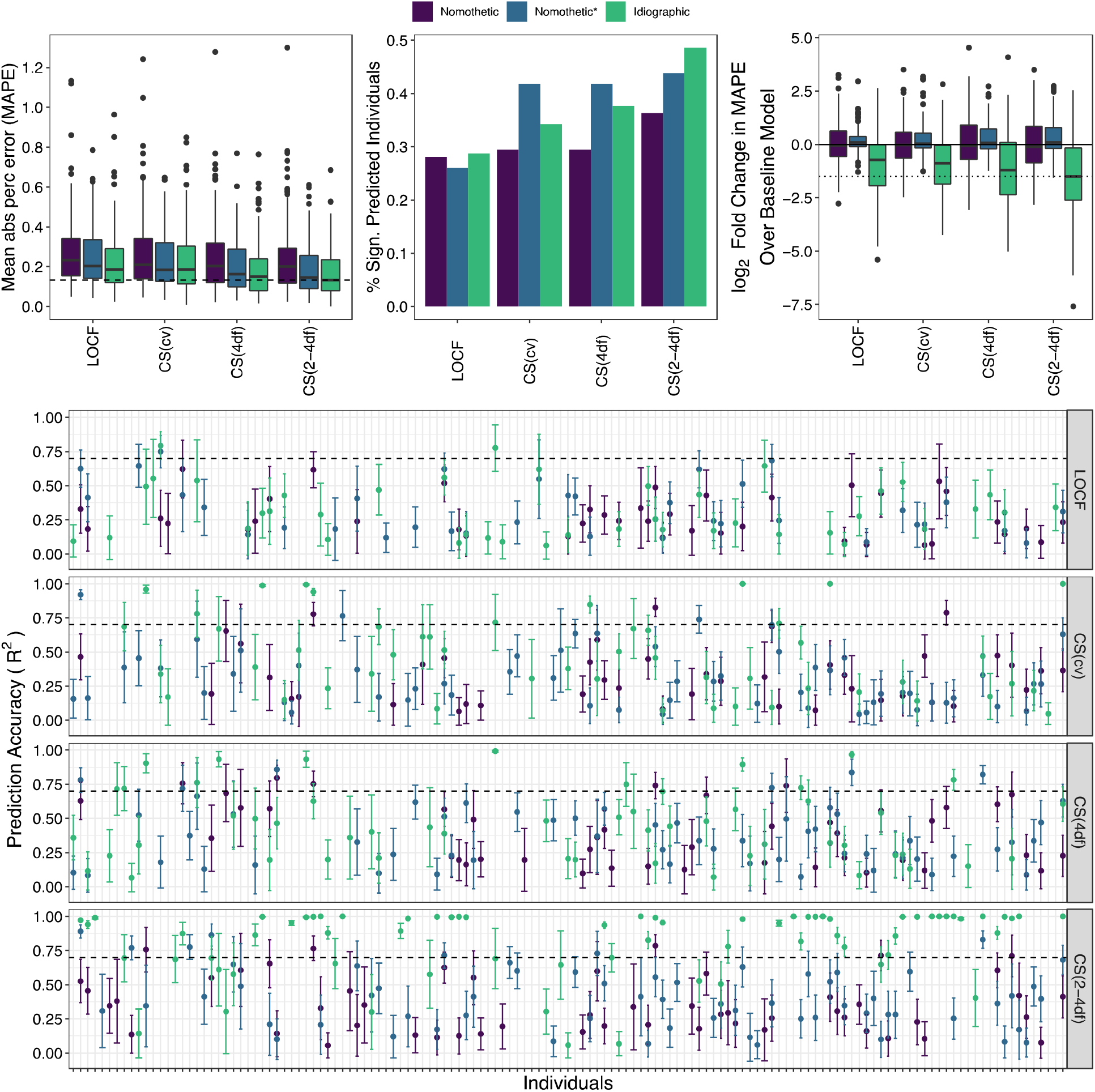
Idiographic models achieve higher individual level prediction accuracy than nomothetic models. **(a)** Box plots of distribution of MAPE across individuals for different models and latent depression traits. The dashed line indicates the median MAPE of the best performing model/latent trait combination, i.e., idiographic model and CS(2-4df) spline. **(b)** Bar plots of the proportion of individuals with significantly predicted mood (R>0 at FDR<5% across individuals) for each latent trait and prediction model. **(c)** Prediction accuracy (R^2^) with 95% CI across all individuals and latent traits. **(d)** Box plot of log2 fold change in CAT-DI prediction accuracy, as measured by MAPE, of feature-based model over the baseline model. Negative log2 fold change in MAPE mean that the feature-based model performs better than the baseline model. All plots are based on individuals with at least five assessments in the test set (N=143). Features were imputed with Autocomplete and CAT-DI was modeled using a logistic elastic net regression. In (a) and (d), the dark black line represents the median value; the box limits show the interquartile range (IQR) from the first (Q1) to third (Q3) quartiles; the whiskers extend to the furthest data point within Q1-1.5*IQR (bottom) and Q3+1.5*IQR (top). LOCF: last observation carried forward. CS(xdf): cubic spline with x degrees of freedom. CS(cv): best-fitting cubic spline according to leave-one-out cross-validation.

Figure 2b illustrates an individual with severe depressive symptoms for whom we can identify a window of disrupted sleep that co-occurred with a clinically significant increase in symptom severity (from mild to severe CAT-DI scores). Subsequently, a return to baseline patterns of sleep coincided with symptom reduction. Quantifying this relationship poses a number of issues, which we turn to next.

### Predicting CAT-DI scores from digital phenotypes

To predict future depression severity scores using digital behavioral phenotypes, we considered three analytical approaches. First, we applied an idiographic approach, whereby we build a separate prediction model for each of the participants. Specifically, for each individual, we train an elastic net regression model using the first 70% of their depression scores and predict the remaining 30% of scores. Second, we applied a nomothetic approach that used data from all participants to build a single model for depression severity prediction using the same analytical steps: we train an elastic net regression model using the first 70% of depression scores of each individual and predict the remaining 30% of scores (Materials and Methods). The result of this nomothetic approach was a single elastic net regression model that makes predictions in all participants.

The main difference between the nomothetic and idiographic approach is that the nomothetic model assumes that each feature has the same relationship with the CAT-DI scores across individuals, for example, that a phone interaction is always associated with an increase in depression score. However, it is possible, and we see this in our data, that an increase in phone interaction can be associated with an increase in symptom severity for one person, but a decrease in another (Figure 2a). The idiographic model allows for this possibility by using a different slope for each feature and individual. In addition, we know that large differences exist in average depression scores between individuals (Figure 1e). To understand the impact of accounting for these differences in a nomothetic approach, we also applied a third approach (referred to as nomothetic*) which includes individual indicator variables in the elastic net regression model in order to allow for potentially different intercepts for each individual. All three models include stay day as a covariate.

To assess whether digital behavioral phenotypes predict mood, we have to deal with the problem that digital phenotypes are acquired daily, while CAT-DI are usually administered every week (and often much less frequently, on average every 10 days). We assume that the CAT-DI indexes a continuously variable trait, but what can we use as the target for our digital predictions when we have such sparsely distributed measures? We can treat this as a problem of imputation, in which case the difficulty reduces to knowing the likely distribution of missing values. However, we also assume that both CAT-DI and digital features only imperfectly reflect a fluctuating latent trait of depression. Thus, our imputation is used not only to fill in missing data points but also to be a closer reflection of the underlying trait that we are trying to predict, namely, depressive severity.

We interpolate the unmeasured estimates of depression by modeling the latent trait as a cubic spline with different degrees of freedom (Figure 3a). For many individuals, CAT-DI values fluctuate considerably during the study, while for others less so. To accommodate this variation, we alter the degrees of freedom of the cubic spline: the more degrees of freedom, the greater the allowed variation. For each individual, we used cubic splines with four degrees of freedom, denoted by CS(4df), degrees of freedom corresponding to the number of observed CAT-DI categories in the training set, denoted by CS(2-4df), and degrees of freedom identified by leave-one-out cross-validation in the training set, denoted by CS(cv). For comparison purposes, we also used a last-observation-carried-forward (LOCF) approach, a naive interpolation method which does not apply any smoothness to the observed trait. In addition, we also include results from analyses done without interpolating CAT-DI but rather modeling the (bi)weekly measurements. Because spline interpolation will cause data leakage across the training-testing split and upwardly bias prediction accuracy, we train our prediction models using cubic spline interpolation on only the training data (first 70% of time series of each individual) and assess prediction accuracy performance in the testing set (last 30%) using the time series generated by applying cubic splines to the entire time series (Figure 3b).

We evaluated the prediction performance of each model and for each latent trait across and within participants. We refer to the former as group level prediction and the later as individual level prediction. Looking at group level prediction performance, compared to within each participant separately, allows us to compute prediction accuracy metrics, e.g., R^2^, as a function of the number of days ahead we are predicting and test for their statistical significance across all predicted observations.

We first evaluated group level prediction accuracy. Figure 4 shows group level prediction performance for each latent trait using the nomothetic, nomothetic*, and idiographic model when the features were imputed with Autocomplete and CAT-DI was modeled using a logistic elastic net regression. We observed that across all latent traits the nomothetic model shows lower prediction accuracy (mean absolute percentage error [MAPE] = 25-28% and R^2^< 5% for all latent traits), compared to the nomothetic* (MAPE = 18-25% and R^2^= 30-46%) or idiographic (MAPE = 16-23% and R^2^=37-66%) models (Figure 4a-b). This is in line with the large proportion of depression scores variance explained by between-individual differences (Figure 1e) which get best captured by the nomothetic* and idiographic models. The idiographic model also showed higher prediction accuracy than the nomothetic(*) model when the features were imputed using softImpute or when CAT-DI was modeled using a linear elastic net regression (Sup Figure 6a-b) as well as when CAT-DI was modeled at the (bi)weekly level without interpolation to get daily level data (Sup Figure 8a-b).

**Figure 6:**
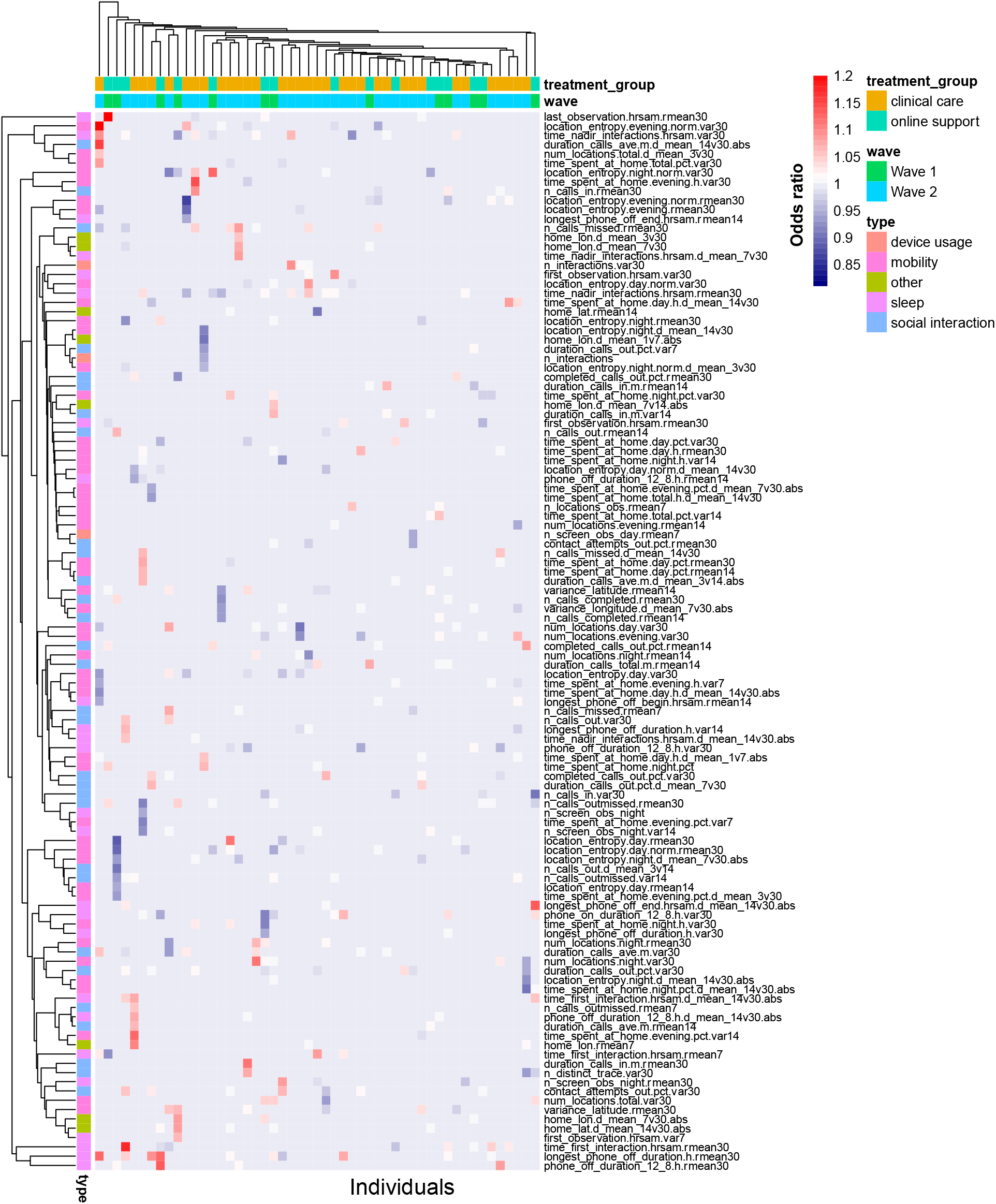
Most predictive behavioral features according to idiographic models. Heatmap of idiographic elastic net regression coefficients for significantly predicted individuals (N=113 with R>0 and FDR<5%). Columns indicate individuals and rows indicate features. For visualization ease, we limit plot to features that have an odds ratio coefficient value above 1.05 or below 0.95 in at least one individual and individuals with at least one feature passing this threshold. The heatmap color indicates the elastic net regularized odds ratio for each feature and individual.

We also compared the prediction performance for each of the different latent traits. As expected, we achieve a higher prediction accuracy for the more highly penalized cubic spline latent traits compared to the LOCF latent trait, as the latter has, by default, a larger amount of variation left to be explained by the features. For example, for the idiographic models, we obtained an R^2^= 66.4% for CS(2-4df) versus 36.9% for LOCF, implying that weekly patterns of depression severity, which are more likely to be captured by the LOCF latent trait, are harder to predict than depression severity patterns over a couple of weeks or months, which are more likely to be captured by the cubic spline latent traits with smallest degrees of freedom.

To understand the effect of time on prediction accuracy, we assessed prediction performance as a function of the number of weeks ahead we are predicting from the last observation in the training set (Figure 4c). The idiographic models achieved high prediction accuracy for depression scores up to three weeks from the last observation in the training set, e.g., R^2^= 84.2% and 73.2% for the CS(2-4df) latent trait to predict observations one week and four weeks ahead, respectively. Prediction accuracy falls below 80% after four weeks.

To quantify the contribution of features on group-level prediction accuracy, we assessed to what extent the features improve the prediction of each model above that achieved by a baseline model that includes just the intercept and study day. Figure 4d-e shows the log2 fold change in CAT-DI prediction accuracy, as measured by MAPE and R^2^, of the feature-based model over the baseline model. The baseline nomothetic model often predicts the same value, i.e., training set intercept, so we cannot compute R^2^. The feature-based idiographic model achieved the greatest improvement in prediction accuracy over the corresponding baseline model, resulting in 65.7% reduction in the MAPE and 7.1% increase in R^2^ over the baseline model for the CS(2-4df) latent trait. The idiographic model also showed higher prediction accuracy than the corresponding baseline model when the features were imputed using softImpute or when CAT-DI was modeled using a linear elastic net regression (Sup Figure 6c-d) as well as when CAT-DI was modeled at the (bi)weekly level (Sup Figure 8c-d). These results suggest that the passive phone features enhance prediction, over and above past CAT-DI and study day, for most individuals in our study.

We next evaluated individual level prediction accuracy (Figure 5). For this analysis, in order to be able to assess the statistical significance of our prediction accuracy within each individual, we only keep individuals with at least five mental health assessments in the test set (N=143). In line with the group level prediction performance, the idiographic model outperformed the other models at the individual level (Figure 5a; median MAPE across individuals for all latent traits = 13.3 - 18.9% versus 20.1-23% for the nomothetic and 14.5-20.4% for the nomothetic* model). Using an idiographic modeling approach, we significantly predicted the future mood for 79.0% of individuals (113 out of 143 with R> 0 and FDR < 5% across individuals) for at least one of the latent traits (Figure 5b), compared to 58.7% and 65.7% of individuals for the nomothetic and nomothetic* model, respectively. The median R^2^ value across significantly predicted individuals for the idiographic models was 47.0% (Figure 5c), compared to 23.7.% and 28.4% for the nomothetic and nomothetic* model, respectively. In addition, for 41.3% of these individuals, the idiographic model had prediction accuracy greater than 70%, demonstrating high predictive power in inferring mood from digital behavioral phenotypes for these individuals, compared to 6.2% and 9.7% for the nomothetic and nomothetic* model, respectively (Figure 5c). The idiographic model also outperformed the nomothetic(*) model when the features were imputed using softImpute or when CAT-DI was modeled using a linear elastic net regression (Sup Figure 7a) as well as when CAT-DI was modeled at the (bi)weekly level (Sup Figure 8e).

**Figure 7:**
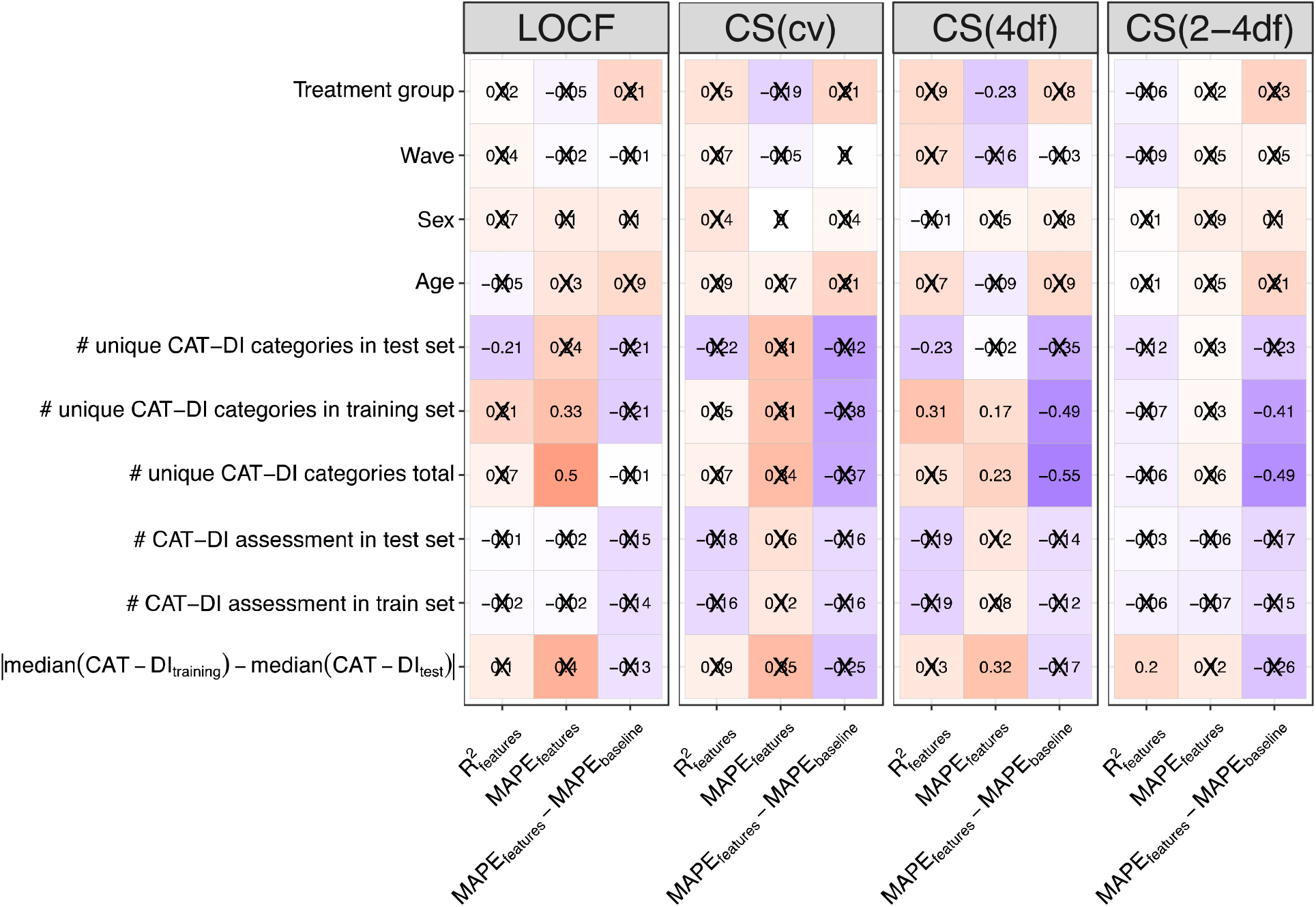
Factors associated with prediction performance of CAT-DI severity scores. Correlation between prediction accuracy of an individual (metrics on the y-axis) and the number of CAT-DI assessment available in the training and test set, the difference in median CAT-DI severity between the training and test set, the number of the unique CAT-DI categories (normal to severe) observed (total and in training and test sets), age, sex, wave, and treatment group (a proxy for depression severity). MAPE: mean absolute percentage error.

Next, we compared individual-level prediction accuracy of each model against the corresponding baseline model that includes just the intercept and study day. Figure 5d and Sup Figure 7c-d show the distribution across individuals of the log2 fold change in CAT-DI prediction accuracy of the feature-based model over the baseline model. In accordance with the group level prediction performance, the feature-based idiographic model achieved the greatest improvement in prediction accuracy over the corresponding baseline model, resulting in a median of over two-fold reduction in the MAPE (Figure 5d; median MAPE of feature-based model across individuals for all latent traits = 13.3 - 18.9% versus 40.1-41.4% for the baseline model). The idiographic model also showed greatest improvement in prediction accuracy over the corresponding baseline model than the nomothetic(*) model when CAT-DI was modeled at the (bi)weekly level (Sup Figure 8f).

To identify the features that most robustly predict depression in each person we extracted top-feature predictors for each individual’s best-fit idiographic model. We limit this analysis to the 113 individuals which showed significant prediction accuracy for at least one of the latent traits. As expected, the study day was predictive of the mood for 63% of individuals and was mainly associated with a decrease in symptom severity (median odds ratio [OR] = 0.86 across individuals). Although no behavioral feature uniformly stood out, as expected by the high correlation between features and heterogeneity in correlation between features and CAT-DI across individuals (Figure 2a), the variation within the last 30 days in the proportion of unique contacts for outgoing texts and messages (a proxy for erratic social behavior), the time of last (first) interaction with the phone after midnight (in the morning) (a proxy for erratic bedtime [wake up time] and sleep quality), and the proportion of time spent at home during the day (a proxy for erratic activity level) were among the top predictors of future mood and were often associated with an increase in symptom severity (OR = 1.05 - 1.23 across features and individuals). The heatmap display of predictor importance in Figure 6 highlights the heterogeneity of passive features for predicting the future across individuals. For example, poor mental health, as indicated by high CAT-DI depression severity scores, was associated with decreased variation in location entropy in the evenings (a proxy for erratic activity level) in the past 30 days for one individual (OR = 0.94) while for another individual it was associated with increased variation (OR =1.20).

### Factors associated with prediction performance

Using digital behavioral features to predict future mood was useful for 74-77% of our cohort and the contribution of the features to the prediction performance varies across these individuals. What might contribute to this variation? Identifying the factors involved might allow us to develop additional models with higher prediction accuracy. To identify factors that are associated with prediction performance, we computed the correlation between accuracy metrics (prediction R^2^ and MAPE of feature-based model and difference in MAPE between feature-based and baseline models) with different study parameters e.g., treatment group, sex, etc. (Figure 7).

Increased variability in depression scores during the study, as measured by the number of unique CAT-DI categories for each individual, were correlated with poorer prediction performance of the feature-based model, as measured by MAPE (Spearman’s ρ= 0.49 and 0.23, p-value = 2.25 x 10^-2^ and 9.79 x 10^-4^ for LOCF and CS(4df) latent traits, respectively). In addition, larger differences in median depression scores between the training and test set for each individual were correlated with poorer prediction performance, as measured by MAPE (Spearman’s ρ=0.32, p-value = 9.11 x 10^-4^ for the CS(4df) latent trait). This suggests that, for some individuals in the study, the training depression scores are higher/lower than the test depression scores (as expected by Sup Figure 4) and that adding the study day or digital phenotypes as a predictor does not completely mediate this issue. The size of the training and test set as well as demographic variables were not strongly correlated to prediction performance.

While we had poorer prediction performance for individuals whose mood shows greater variability during the course of the study, these are also the individuals for which using a feature-based model improves prediction accuracy compared to a baseline model that predicts based on past depression severity and study day alone. Specifically, larger variability in depression scores for each individual was correlated with better prediction performance of a feature-based model than a baseline model, as measured by difference in MAPE between the two models (Spearman’s ρ=-0.54 and −0.49, p-value = 5.96x 10^-4^ and p-value = 4.46 x 10^-3^ for the CS(4df) and CS(2-4df) latent traits, respectively).

## Discussion

In this paper, we showed the feasibility of longitudinally measuring depressive symptoms over 183 individuals for up to 10 months using computerized adaptive testing and passively and continuously measuring behavioral data captured from the sensors built into smartphones. Using a combination of cubic spline interpolation and idiographic prediction models, we were able to impute and predict a latent depression trait on a hold-out set of each individual several weeks in advance.

Our ability to longitudinally assess depressive symptoms and behavior within many individuals and over a long period of time enabled us to assess how far out we can predict depressive symptoms, how variable prediction accuracy can be across different individuals, and what factors contribute to this variability. In addition, it enabled us to assess the contribution of behavioral features to prediction accuracy above and beyond that of prior symptom severity or study day alone. We observed that prediction accuracy dropped below 70% after four weeks. In addition, prediction accuracy varied considerably across individuals as did the contribution of the features to this accuracy. Individuals with large variability in symptom severity during the course of the study (such as those in clinical care) were harder to predict but benefited the most from using behavioral features. We expect that pairing digital phenotypes from smartphones with behavioral phenotypes from wearable devices, which are worn continuously and might measure behavior with less error, as well as addition of phenotypes, like those from electronic health records, could help address some of these challenges.

Our results are consistent with other studies that predict daily mood as measured by ecological momentary assessments or a short screener (i.e., PHQ2^17^) and confirm the superior prediction performance of idiographic models over nomothetic ones. Our study goes further, by exploring if the superior prediction accuracy of idiographic models is a result of better modeling the relationship between features and mood or simply of better modeling the baseline mood of each individual. We show that a large part of the increase in prediction performance of idiographic models is due to the latter, as indicated by the increase in prediction performance between the nomothetic and modified nomothetic models.

High-burden studies over long time periods may result in drop-out, particularly for depressed individuals^33^. In our case, we observed that attrition for CAT-DI assessment was linear over the follow-up period, except for the first two weeks during which a large proportion of individuals which received online support dropped out (typical of online mental health studies^34^). In addition, participants which received clinical care were more adherent than those which received online support, despite endorsing more severe depressive symptoms. These participants had regular in-person treatment sessions during which they were instructed to complete any missing assessments emphasizing the importance of using reminders or incentives for online mental health studies.

There are several limitations in the current study. First, the idiographic models that we use here are fit separately for each individual and might not thus maximize statistical power. In addition, they assume a (log-)linear relationship between behavioral features and depression severity and will fit poorly if this assumption is violated. One potential alternative is to employ mixed models that jointly model data from all individuals using individual-specific slopes and low degree polynomials. However, due to the high dimensionality of our data, such models are hard to implement. Second, while it is well established that Computerized Adaptive Testing can be repeatedly administration to the same person over time without response set bias due to adaptive question sets^25^, extended use over months might still lead to limited response bias^35^. Third, the adaptive nature of CAT-DI, which might assess different symptoms for different individuals, frustrates joint analyses. Fourth, the imputation method used for imputing digital behavioral features assumes data to be missing at random (MAR), meaning missingness depended on observed data^36^. While this assumption is hard to test, MAR seems quite plausible in our study given that the data is missing more often for participants that did not receive regular reminders. In addition, research has shown that violation of the MAR assumption does not seriously distort parameter estimates^37^. Finally, the age and gender distribution in our participants may limit the generalizability of our findings to the wider population.

In conclusion, our study verified the feasibility of using passively collected digital behavioral phenotypes from smartphones to predict depressive symptoms weeks in advance. Its key novelty lies in the use of computerized adaptive testing, which enabled us to obtain high-quality longitudinal assessments of mood on 183 individuals over many months, and in the use of personalized prediction models, which offer a much higher predictive power compared to nomothetic models. Ultimately, we expect that the method will lead to a screening and detection system that will alert clinicians in real-time to initiate or adapt treatment as required. Moreover, as passive phenotyping becomes more scalable for hundreds of thousands of individuals, we expected that this method will enable large genome and phenome-wide association studies for psychiatric genetic research.

## Materials and Methods

### Study participants and treatment protocol

Participants are University of California Los Angeles (UCLA) students experiencing mild to severe symptoms of depression or anxiety enrolled as part of the STAND program^29^ developed under the UCLA Depression Grand Challenge^38^ treatment arm. All UCLA students aged 18 or older who had internet access and were fluent in English were eligible to participate. STAND enrolled participants in two waves. The first wave enrolled participants from April 2017 to June 2018. The second wave of enrollment began at the start of the academic year in 2018 and continued for three years, during which time, from March 2020, a Safer-At-Home order was imposed in Los Angeles to control the spread of COVID-19. All participants are offered behavioral health tracking through the AWARE^27^ framework and had to install the app in order to be included in the study. All participants provided written informed consent for the study protocol approved by the UCLA institutional review board (IRB #16-001395 for those receiving online support and #17-001365 for those receiving clinical support).

Depression symptom severity at baseline and during the course of the study was assessed using the Computerized Adaptive Testing Depression Inventory^25^ (CAT-DI), a validated online mental health tracker. Computerized adaptive testing is a technology for interactive administration of tests that tailors the test to the patient^26^. Tests are ‘adaptive’ in the sense that the testing is driven by an algorithm that selects questions in real-time and in response to the ongoing responses of the patient. CAT-DI uses item response theory to select a small number of questions from a large bank, thus providing a powerful and efficient way to detect psychiatric illness without suffering response fatigue.

Participants were classified into treatment groups based on their depression and anxiety scores at baseline, which indicated the severity of symptoms in those domains. Individuals who are not currently experiencing symptoms of depression (CAT-DI score < 35) or anxiety are offered the opportunity to participate in the study without an active treatment component by contributing CAT-DI assessment. These individuals are excluded from our analyses as they do not show any variation in CAT-DI. Participants that exhibited scores below the moderate depression range (CAT-DI score < 74) were offered internet-based cognitive behavioral therapy, which includes adjunctive support provided by trained peers or clinical psychology graduate students via video chat or in person. Eligible participants with symptoms in this range were excluded if they were currently receiving cognitive behavioral therapy, refused to install the AWARE phone sensor app, or were planning an extended absence during the intervention period. Participants that exhibited scores in the range of severe depression symptoms (CAT-DI score 75-100) or who endorsed current suicidality were offered in-person clinical care which included evidence-based psychological treatment with option for medication management. Additional exclusion criteria were applied to participants with symptoms in this range, which included clinically-assessed severe psychopathology requiring intensive treatment, multiple recent suicide attempts resulting in hospitalization, or significant psychotic symptoms unrelated to major depressive or bipolar manic episodes. These criteria were determined through further clinical assessment. Participants with symptoms in this range were also excluded if they were unwilling to provide a blood sample or transfer care to the study team while receiving treatment in the STAND program.

Depression symptom severity was assessed up to every other week for the participants that received online support (both waves), i.e., those with mild to moderate symptoms, and every week for the participants that received in-person clinical care, i.e., those with severe symptoms (Sup Figure 2a). Participants that received in-person care had also four in-person assessment events, at weeks 8, 16, 28, and 40, prior to the COVID-19 pandemic. Thus, Wave 1 participants can have a maximum of 13 CAT-DI assessments while Wave 2 participants can have a maximum of 21 (online support) or 44 assessments, depending on severity and excluding initial assessments prior to treatment assignment.

CAT-DI was assessed at least one time for 437 individuals that installed the AWARE app. Here, we limit our prediction analyses to individuals that have at least five CAT-DI assessments (N=238; since we need at least four points to interpolate CAT-DI in the training set), have at least 60 days of sensor data in the same period for which CAT-DI data is also available (N=189), and show variation in their CAT-DI scores in the training set (N=183), which is necessary in order to build prediction models.

### Adherence to CAT-DI assessment protocol and factors affecting adherence

To assess if participant adherence to CAT-DI assessments varied across enrollment waves and treatment groups, we used a logistic regression with the proportion of CAT-DI assessments a participant completed as the dependent variable and the enrollment waves or treatment groups as independent variables. A similar model was used to assess impact of sex and age on participant adherence (results presented in the Supplement). To assess if participant adherence varied with time in the study, we used a logistic regression random effect model, as implemented in the lmerTest^39^ R package, with an indicator variable for the individual remining in the study for each required assessment as the dependent variable and a continuous study week as an independent variable. An individual-specific random effect was used to account for repeated measurement of each individual during the study. A likelihood ratio test was used to test for the significant of the effect of each independent variable against the appropriate null model.

### Variance partition of CAT-DI metrics

We calculate the proportion of CAT-DI severity variance explained by different study parameters using a linear mixed model as implemented in the R package variancePartition^40^ with the subject id, study id, season, sex, and year modeled as random variables while the day of the study, the age of the subject, and a binary variable indicating the dates before or after the safer at home order was issued in California modeled as fixed, i.e., 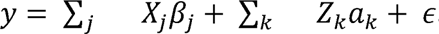, where *y* is the vector of the CAT-DI values across all subjects and time points, *X_j_* is the matrix of j^th^ fixed effect with coefficients *β_j_*, *Z_k_* is the matrix corresponding to the k^th^ random effect with coefficients *a_k_* drawn from a normal distribution with variance 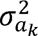. The noise term, *ε*, is drawn from a normal distribution with variance 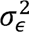. All parameters are estimated with maximum likelihood^42^. Variance terms for the fixed effects are computed using the post hoc calculation 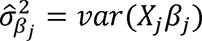. The total variance is 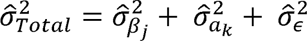 so that the fraction of variance explained by the j^th^ fixed effect is 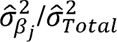, by the k^th^ random effect is 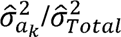, and the residual variance is 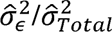. Confidence intervals for variance explained were calculated using parametric bootstrap sampling as implemented in the R package variancePartition^41^.

### Feature extraction from smartphone sensors

We describe feature extraction in detail in below. Broadly, we extracted 23 features related to mobility, e.g., location entropy, 13 related to sleep and circadian rhythm, e.g., hours of uninterrupted sleep, 18 related to social interaction, e.g., duration of outgoing calls, and two related to mobile device usage, e.g., number of interactions with phone per day. Each of these features was calculated on a daily basis. Furthermore, each of these features was computed over three daily non-overlapping time windows of equal duration (night 00:00-08:00, day 08:00-16:00, evening 16:00-00:00), under the hypothesis that participant behavior may be more or less variable based on external constraints such as a regular class schedule during daytime hours.

In addition, considering a participant’s current mental state may be influenced by patterns of behavior from days prior, sliding window averages of each of the daily features were calculated over multiple sliding windows ranging from three days to one month prior to the current day, i.e., windows of length three, seven, 14, and 30 days. The variance of each feature was also calculated over these same windows, to estimate whether behavior had been stable or variable during that time, e.g., were there large fluctuations in sleep time over the past week?

Finally, under the hypothesis that recent *changes* in behavior may be more indicative of changes in mental state than absolute measures, a final set of transformations were applied to each feature. These transformations compared the sliding window means of two different durations against each other, to estimate the change in behavior during one window over that of a longer duration window (the longer window serving as a local baseline for the participant). This allowed estimates from the raw features of whether, e.g., the participant had slept less last night than typical over the past week or slept less on average in the last week than typical over the last month. All of these transformations were applied to the base features extracted from sensor data and included as separate features fed into subsequent regression approaches.

In total, 1,325 raw and transformed features were extracted and included in the final analysis.

#### Preprocessing features

Each sensor collected through the AWARE framework is stored separately with a common set of data items (device identifier, timestamp, etc.) as well as a set of items unique to each sensor (sensor-specific items such as GPS coordinates, screen state, etc.). Data from each sensor was preprocessed to convert Unix UTC timestamps into local time, remove duplicate logging entries, and remove entries with missing sensor data. Additionally, some data labels that are numerically coded during data collection (e.g., screen state) were converted to human-readable labels for ease of interpretation.

#### Mobility features

Location data was divided into 24-hour windows starting and ending at midnight each day. To identify locations where participants spent time, GPS data were filtered to identify observations where the participants were stationary since the previous observation. Stationary observations were those defined as having an average speed of <0.7 meters per second (approximately half the average walking speed of the average adult). These stationary observations were then clustered using hierarchical clustering to identify unique locations in which participants spent time during each day. Hierarchical clustering was chosen over k-means and density-based approaches such as DBSCAN due to its ability to deterministically assign clusters to locations with a precisely defined and consistent radius, independent of occasional data missingness.

Locations were defined to have a maximum radius of 400 m, a sufficient radius to account for noise in GPS observations. Clusters were then filtered to exclude any location in which the participant spent less than 15 minutes over the day to exclude location artifacts, e.g., a participant being stuck in traffic during daily commute, or passing through the same area of campus multiple times in a day. To address data missingness in situations where GPS observations were not received at regular intervals, locations were linearly interpolated to provide an estimated location every 3 minutes.

For each day, a home location was assigned based on the location each participant spent the most time in between the hours of midnight to eight am. This approach allowed for better interpretation of behavior for participants who split time between multiple living situations, for example, students who return home for the weekend or a vacation. Next, multiple features were extracted from this location data, including total time spent at home each day, total number of locations visited, overall location entropy, and normalized location entropy. Each of these features was additionally computed over three daily non-overlapping time windows of equal duration (night 00:00-08:00, day 08:00-16:00, evening 16:00-00:00), under the hypothesis that participant behavior may be more or less variable based on external constraints such as a regular class schedule during daytime hours.

#### Sleep and circadian rhythm features

Sleep and circadian rhythm features were extracted from logs of participant interactions with their phone, following prior work showing that last interaction with the phone at night can serve as a reasonable proxy for bedtime, and first interaction in the morning for waketime. The longest phone-off period (or assumed uninterrupted sleep duration) was tracked each night, as well as the beginning and end time of that window as estimates of bedtime and waketime. To account for participants who may have interrupted sleep, the time spent using the phone between the hours of midnight and 8 am was also tracked to account for participants who may use their phone briefly in the middle of the night but are otherwise asleep for the majority of that window. Finally, time-varying kernel density estimates were derived using the total set of phone interactions, to estimate the daily time nadir of interactions, as an additional proxy for the time of overall circadian digital activity nadir.

#### Social interaction and other device usage features

Additional social interaction features were extracted from anonymized logs of participant calls and text messages sent and received from their smartphone device. Features extracted from this data include, for example, the total number of phone calls made, total time spent on the phone, and percentage of calls connected that were outgoing (i.e., dialed by the participant) versus incoming. Due to OS restrictions, sensors needed to extract text message features are not available on iOS devices and were only computed for the 15 participants with Android devices.

### Imputation of smartphone-based features

To address the missing features problem (Sup Figure 4), we considered two different imputation methods: matrix completion via iterative soft-thresholder SVD, as implemented in the R package softImpute, and AutoComplete, a deep-learning imputation method that employs copy-masking to propagate missingness patterns present in the data. Both approaches were applied separately to each individual as follows. First, we removed features that exhibited > 90% missingness for that individual. Next, we trained the imputation model on the training split alone. Finally, each imputation model was applied to the training and test dataset to impute the features for that individual. Before prediction, we normalize all features to have zero mean and unit standard deviation using mean and standard deviation estimates from the training set alone.

### Mapping of behavioral features to DSM-5 Major Depressive Disorder criteria

The set of features described above map onto only a subset of DSM criteria that are closely associated with externally observable behaviors (Sup Figure 5) - sleep, loss of energy, and anhedonia (to the extent it is severe enough to globally reduce self-initiated activity). Other DSM criteria such as weight change, appetite disturbance, and psychomotor agitation/retardation are in theory also directly observable, but less so with the set of sensors available on a standard smartphone. For these criteria, other device sensors - for instance, smartwatch sensors - may be more applicable in the detection of e.g., fidgeting associated with psychomotor agitation. A final set of DSM criteria include those primarily subjective findings - depressed mood, feelings of worthlessness, suicidal ideation - which inherently require self-report to directly assess. Given that only 5 of 9 criteria are required for the diagnosis of MDD, an individual patient’s set of symptoms may overlap minimally with those symptoms we expect to measure with the features described above. However, for others, the above features may cover a more significant portion of their symptom presentation and do a better job directly quantifying fluctuations in DSM-5 criteria for that individual.

### Imputation of CAT-DI severity scores for prediction models

To get daily-level CAT-DI severity scores, we interpolate the scores for each individual across the whole time series (ground truth) or only the time series corresponding to the training set (70% of the time series) by moving the last CAT-DI score forward, denoted by LOCF, or by smoothing the CAT-DI scores using cubic splines with different degrees of freedom (Figure 3a). Cubic smoothing spline fitting was done using the *smooth.spline* function from the *stats* package in R. We consider cubic splines with four degrees of freedom (denoted by CS(4df) and corresponding to the number of possible CAT-DI severity categories, i.e. normal, mild, moderate, and severe), cubic splines with degrees of freedom equal to the number of observed CAT-DI categories for each individual in the training set (ranging from two to four and denoted by CS(2-4df)), and degrees of freedom identified by ordinary leave-one-out cross-validation in the training set (denoted by CS(cv)).

### Nomothetic and idiographic prediction models of future mood

We split the data for each individual into a training (70% of trajectory) and a test set (remaining 30% of trajectory). To predict the future mood of each individual in the test set from smartphone-based features in the test set, we train an elastic net logistic or linear regression model^41^ in the train set. We set a, i.e., the mixing parameter between ridge regression and lasso, to 0.5 and use 10-fold cross-validation to find the value for parameter A, i.e., the shrinkage parameter. For the idiographic models, we train separate elastic net models for each individual while for the nomothetic and modified nomothetic models we train one model across all individuals. To account for individual differences in the average CAT-DI severity scores in the training set, the modified nomothetic model fits individual-specific intercepts by including individual indicator variables in the regression model. This is similar in nature to a random intercept mixed model where each individual has their own intercept. Note that the test data are the same for all of these models, i.e., the remaining 30% of each individual’s trajectories. Predictions outside the CAT-DI severity range, i.e., [0,100], are set to NA and not considered for model evaluation. We compute prediction accuracy metrics by computing the Pearson’s product-moment correlation coefficient (R) between observed and predicted depression scores in the test set across and within individuals as well as the squared Pearson coefficient (R^2^). To assess the significance of the prediction accuracy we use a one-sided paired test for Pearson’s product-moment correlation coefficient, as implemented in the *cor.test* function of the stats^42^ R package, and a likelihood ratio test for the significance of R^2^. We use the Benjamini-Hochberg procedure^43^ to control the false discovery rate across individuals at 5%.

## Data Availability

The datasets generated and analyzed during the current study are available from the corresponding author upon reasonable request.

## Code Availability

The code that supports the findings of this study is available online at https://github.com/BrunildaBalliu/stand_mood_prediction.

## Data Availability

All data produced in the present study are available upon reasonable request to the authors.

## Acknowledgments

The authors gratefully acknowledge all study participants. S.S was funded in part by NIH grant R35GM125055 and NSF grants III-1705121 and CAREER-1943497. D.S. was supported by NSF-NRT #1829071. J.F. was funded by NIH grant R01MH122569.

## Author Contributions

BB and JF conceived of the project. D.C., V.C., and J.K. participated in the subject recruitment and data collection. BB lead the data analysis with contributions from CD, LS, AW, and DS. BB and JF wrote the first draft of the manuscript with contribution from CD and SS. All authors contributed to subsequent edits of the manuscript and approved the final manuscript.

