## Supplemental Figures for "Personalized Mood Prediction from Patterns of Behavior Collected with Smartphones"

### *Supplemental Material*

Brunilda Balliu*[1,2,3], Chris Douglas[4], Darsol Seok[5], Liat Shenhav[6], Yue Wu[6], Doxa Chatzopoulou[5], William Kaiser[7], Victor Chen[7], Jennifer Kim[5], Sandeep Deverasetty[5], Inna Arnaudova[5], Robert Gibbons[8], Eliza Congdon[4,5], Michelle G. Craske[4,9], Nelson Freimer[4,10], Eran Halperin[6], Sriram Sankararaman[1,6,10], Jonathan Flint[4,10]*

[1]Departments of Computational Medicine, University of California Los Angeles, Los Angeles, USA,

[2]Departments of Pathology and Laboratory Medicine, University of California Los Angeles, Los Angeles, USA,

[3]Department of Biostatistics, University of California Los Angeles, Los Angeles, USA,

[4]Department of Psychiatry and Biobehavioral Science, University of California Los Angeles, Los Angeles, USA,

[5]Semel Institute for Neuroscience and Human Behavior, University of California Los Angeles, Los Angeles, USA,

[6]Department of Computer Science, University of California Los Angeles, Los Angeles, USA,

[7]Department of Electrical Engineering, University of California Los Angeles, Los Angeles, USA,

[8]Departments of Medicine, Public Health Sciences and Comparative Human Development, University of Chicago, USA

[9]Department of Psychology, University of California Los Angeles, Los Angeles, USA,

[10]Department of Human Genetics, University of California Los Angeles, Los Angeles, USA

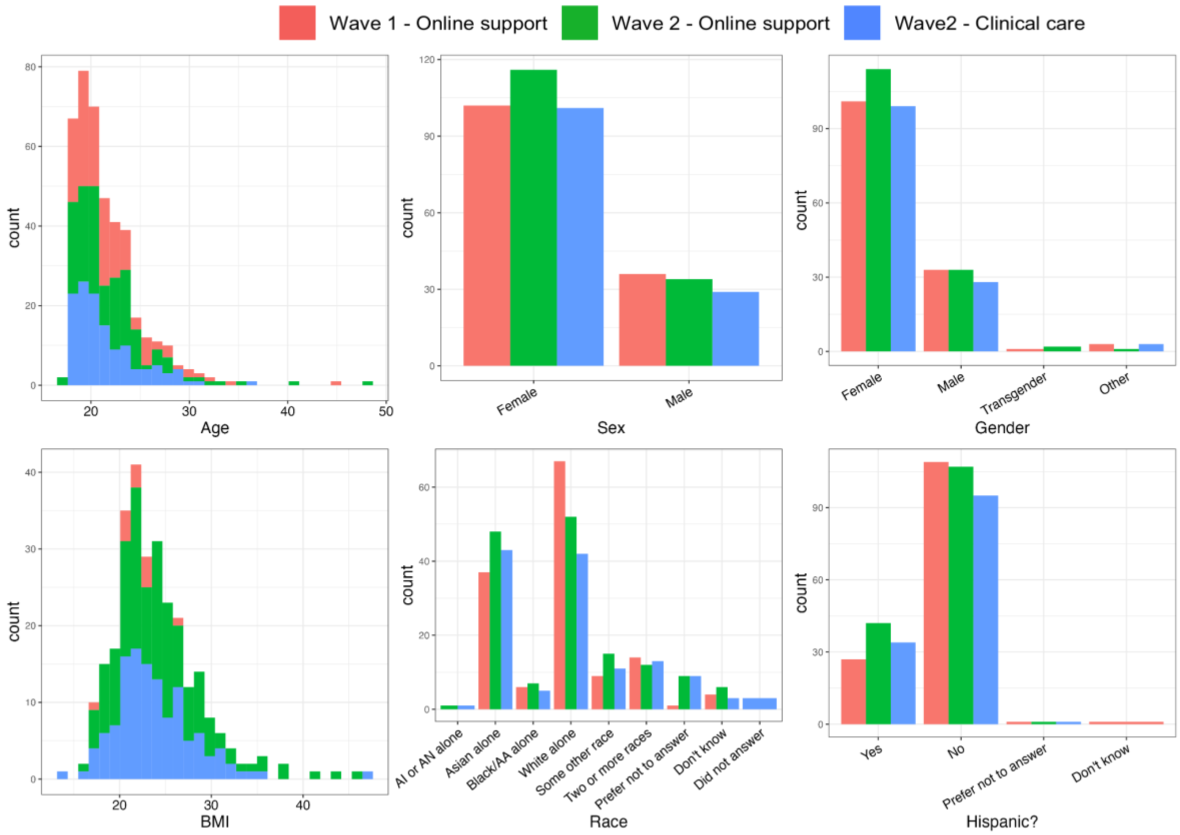


*Sup Figure 1:* ***Demographic information for participants in each wave****. First row: histogram of age and bar plot of sex and gender. Second row: histogram of BMI and bar plot of race and ethnicity. Color indicates wave and treatment protocol combination. AI or AN: American Indian or Alaska Native. AA: African American.*

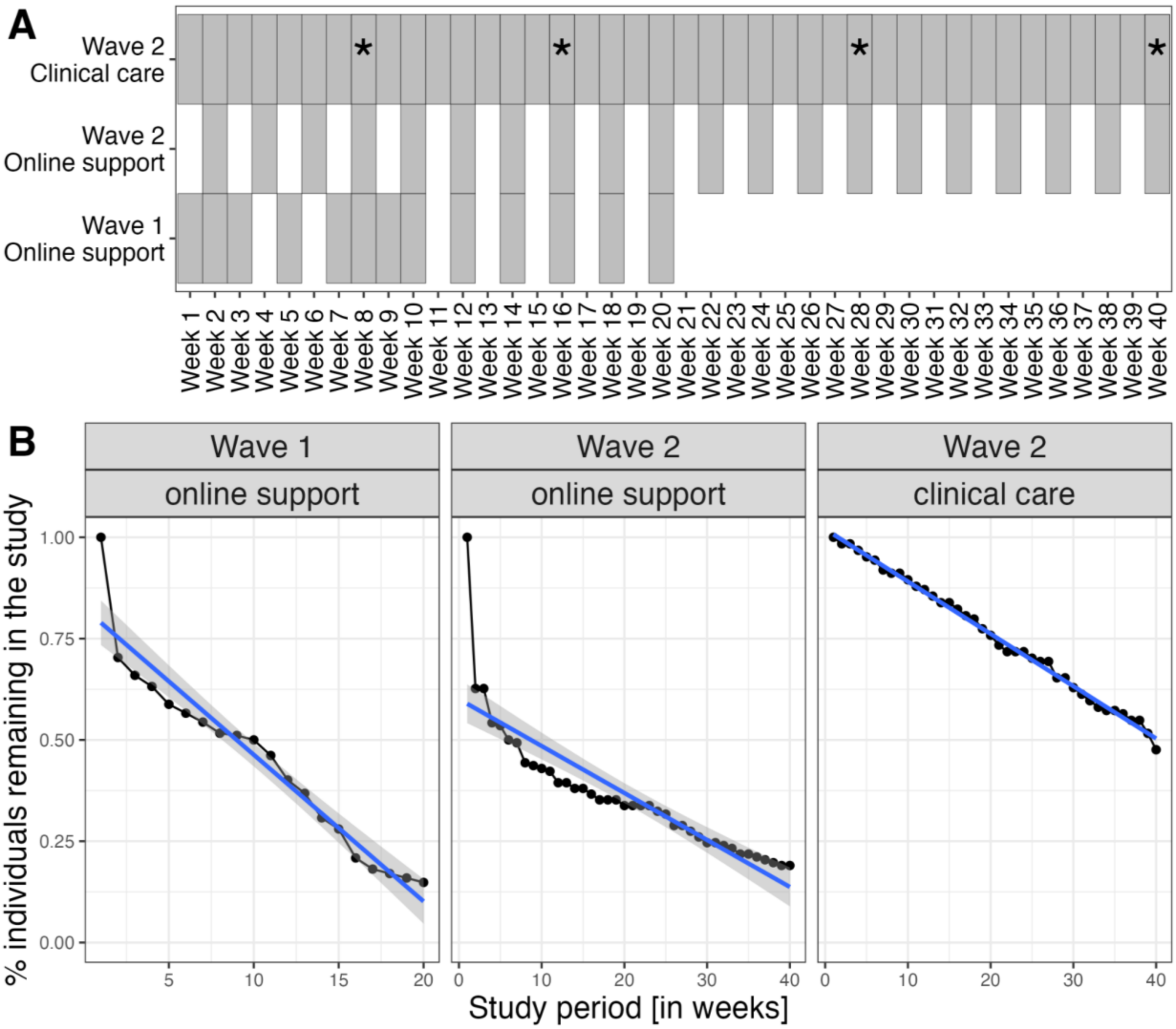


*Sup Figure 2:* ***CAT-DI administration protocol and compliance with CAT-DI assessment protocol for each wave and treatment group. (A)*** *CAT-DI administration schedule. Each box indicates a week during which participants in each group were expected to complete the CAT-DI. Asterisks indicate weeks with additional in-person administrations of CAT-DI for Wave 2 participants which received clinical care. (B) Participant CAT-DI retention rate for each enrollment wave and treatment group. The x-axis shows weeks from the beginning of the study for each participant while the y axis shows the proportion of individuals that were still completing the CAT-DI at that week. The continuous lines show the linear regression fit with 95% confidence intervals (gray shading).*

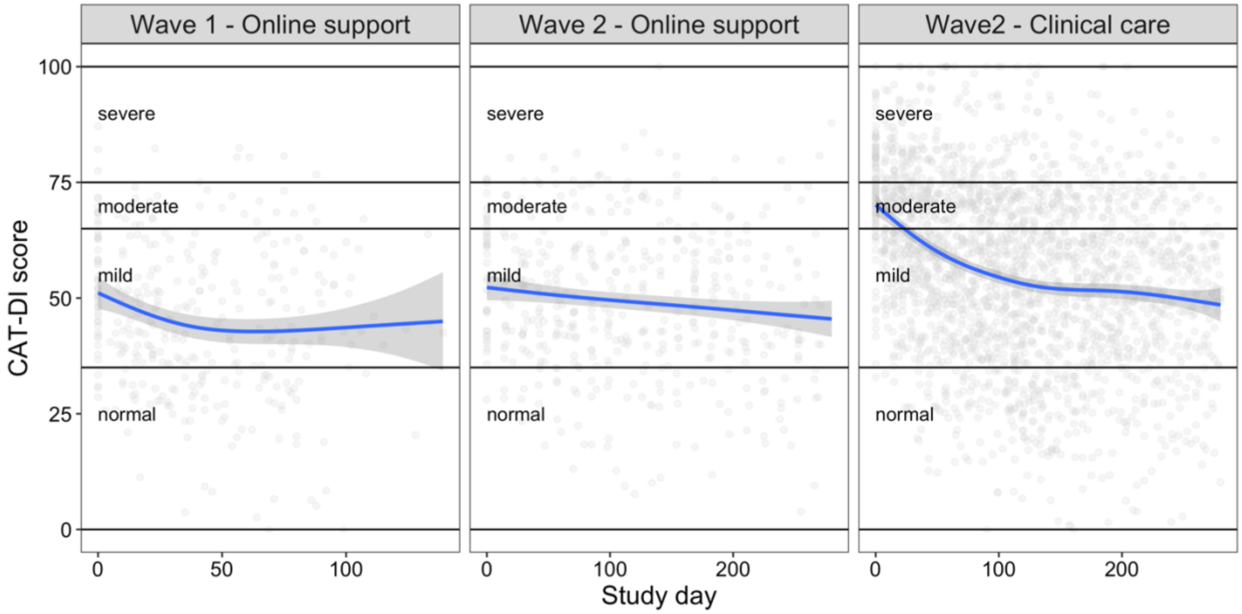


*Sup Figure 3:* ***Effect of therapy per wave and treatment group.*** *The x-axis shows the study day with zero indicating the first day of CAT-DI assessment for each individual. The y-axis indicates the CAT-DI severity score for each individual / day in the study. The blue line indicates the fit of a generalized additive model with y ~ s(day + wave: treatment group, bs = "cs") and gaussian family.*

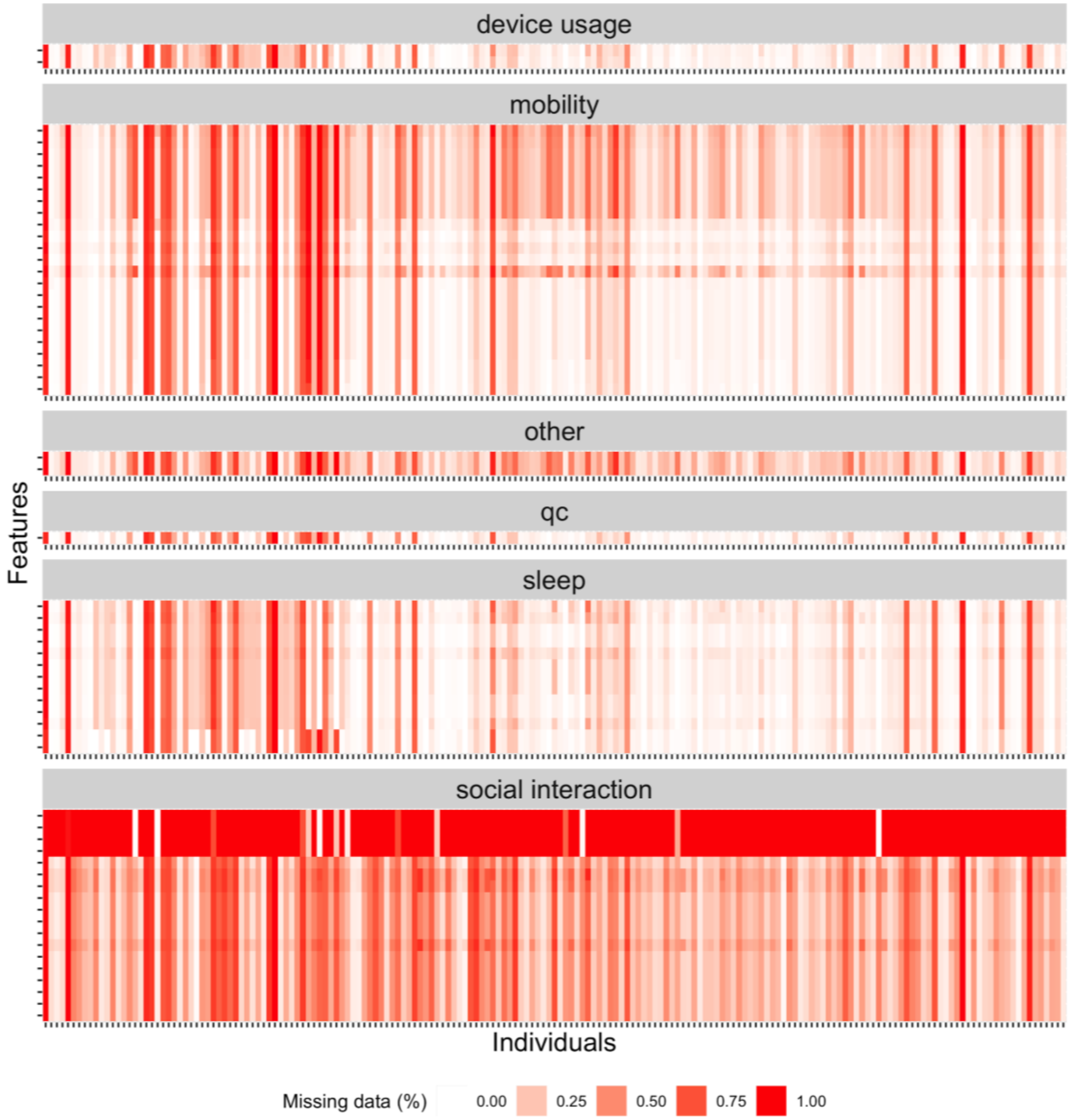


*Sup Figure 4:* ***Missing feature data summary.*** *Heat map showing missing data percentage in each of the four types of features extracted from smartphone data for all individuals. Each tick on the x-axis (y-axis) represents an individual (feature). For ease of plotting, we have excluded transformation-based features. For participants with iOS devices (majority of individuals), we did not have any information on social interaction features related to text message information due to permission. These features are excluded from analyses when considering individuals with iOS devices.*

| | | Depressed mood | Diminished interest or loss of pleasure activities (anhedonia) | Weight change or appetite disturbance | Sleep disturbance | Psycho motor agitation or retardation | Fatigue or loss of energy | Feelings of worthlessness | Diminished concentration; indecisiveness | Suicidal ideation/intent |
|---|---|---|---|---|---|---|---|---|---|---|
| **Mobility features** | *Variance in latitude* | | • | | | | | | | |
| | *Variance in longitude* | | • | | | | | | | |
| | *Number of locations visited in total* | | • | | | | | | | |
| | *Number of locations visited at night* | | • | | | | | | | |
| | *Number of locations visited during the day* | | • | | | | | | | |
| | *Number of locations visited in the evening* | | • | | | | | | | |
| | *Location entropy over full day* | | • | | | | | | | |
| | *Location entropy over full day (normalized)* | | • | | | | | | | |
| | *Location entropy at night* | | • | | | | | | | |
| | *Location entropy at night (normalized)* | | • | | | | | | | |
| | *Location entropy during the day* | | • | | | | | | | |
| | *Location entropy during the day (normalized)* | | • | | | | | | | |
| | *Location entropy in the evening* | | • | | | | | | | |
| | *Location entropy in the evening (normalized)* | | • | | | | | | | |
| | *Time spent at home total (hours)* | | • | | | | • | | | |
| | *Time spent at home at night (hours)* | | • | | | | • | | | |
| | *Time spent at home during the day (hours)* | | • | | | | • | | | |
| | *Time spent at home in the evening (hours)* | | • | | | | • | | | |
| | *Percentage of time spent at home in total (%)* | | • | | | | • | | | |
| | *Percentage of time spent at home at night (%)* | | • | | | | • | | | |
| | *Percentage of time spent at home during the day (%)* | | • | | | | • | | | |
| | *Percentage of time spent at home in the evening (%)* | | • | | | | • | | | |
| **Phone interactions** | *Number of phone interactions during day* | | | | | | | | • | |
| | *Number of phone interactions at night* | | | | • | | | | | |
| **Sleep** | *Duration of longest phone off period (hours)* | | | | • | | | | | |
| | *Beginning of longest phone off period* | | | | • | | | | | |
| | *End of longest phone off period* | | | | • | | | | | |
| | *Phone on duration midnight to 8am (hours)* | | | | • | | | | | |
| | *Phone off duration midnight to 8am (hours)* | | | | • | | | | | |
| | *Percentage of time phone on midnight to 8am (%)* | | | | • | | | | | |
| | *Time of nadir of phone interactions* | | | | • | | | | | |
| **Social interaction** | *Total number of calls* | | • | | | | | | | |
| | *Number of incoming calls attempted* | | • | | | | | | | |
| | *Number of outgoing calls attempted* | | • | | | | | | | |
| | *Number of completed calls* | | • | | | | | | | |
| | *Number of missed calls* | | • | | | | | | | |
| | *Number of unanswered outgoing calls* | | • | | | | | | | |
| | *Percentage of completed outgoing calls (%)* | | • | | | | | | | |
| | *Percentage of unanswered outgoing calls (%)* | | • | | | | | | | |
| | *Duration of incoming calls (minutes)* | | • | | | | | | | |
| | *Duration of outgoing calls (minutes)* | | • | | | | | | | |
| | *Total time spent on phone (minutes)* | | • | | | | | | | |
| | *Average duration per call (minutes)* | | • | | | | | | | |
| | *Duration of outgoing calls (%)* | | • | | | | | | | |
| | *Number of distinct message contacts* | | • | | | | | | | |
| | *Total number of messages* | | • | | | | | | | |
| | *Number of incoming messages* | | • | | | | | | | |
| | *Number of outgoing messages* | | • | | | | | | | |
| | *Percent of messages outgoing* | | • | | | | | | | |

*Sup Figure 5: **Mapping of sensor-derived behavioral features to DSM5 Major Depressive Disorder criteria.** The individual behavioral features derived from phone sensors map primarily to the DSM criteria of disrupted sleep, loss of energy, and anhedonia. Each of these base features is further transformed to look for deviations from individual baseline over varying time scales (e.g., last day's deviation from the weekly average) to arrive at the final set of behavioral features.*

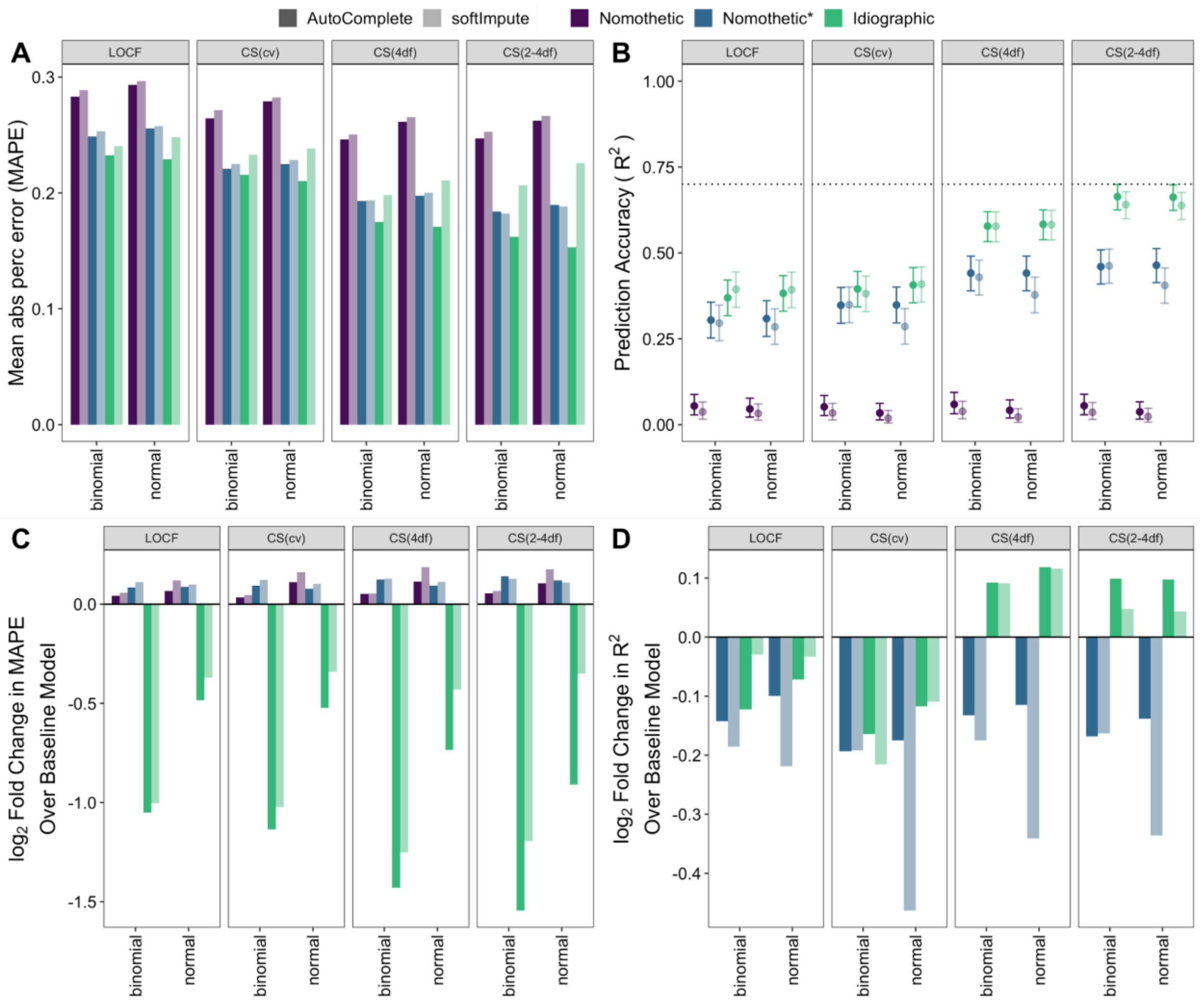


Sup Figure 6: **Idiographic models achieve higher group level prediction accuracy than nomothetic models. (A-B)** CAT-DI prediction accuracy across all individuals in the test set as measured by MAPE (A) and $R^2$ (B) across all individuals for different latent depression traits (panel), modeling approaches (color), CAT-DI regression model (x-axis), and feature imputation methods (transparency). The dotted line in B indicates 70% prediction accuracy and bars indicate 95% confidence intervals of $R^2$. **(C-D)** log2 fold change in CAT-DI prediction accuracy, as measured by MAPE (C) and $R^2$ (D), of feature-based model over the baseline model for different latent depression traits, modeling approaches, CAT-DI regression models, and feature imputation methods. Negative log2 fold change in MAPE and positive log2 fold change in $R^2$ mean that the feature-based model performs better than the baseline model. A log2 fold change in MAPE of -1 means that the prediction error of the baseline model is twice as large as that of the feature based model. MAPE: mean absolute percent error. LOCF: last observation carried forward. CS(xdf): cubic spline with x degrees of freedom. CS(cv): best-fitting cubic spline according to leave-one-out cross-validation.

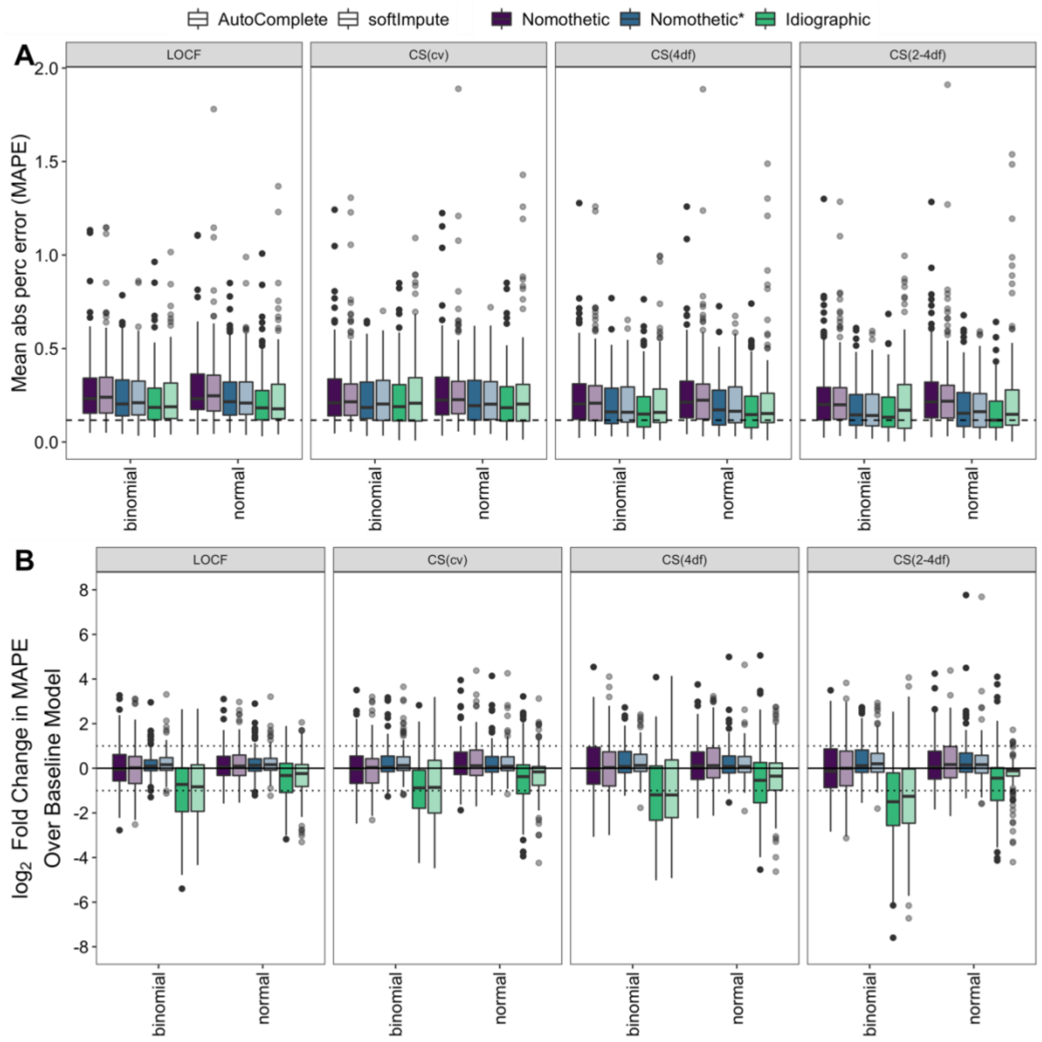


*Sup Figure 7:* ***Idiographic models achieve higher individual-level prediction accuracy than nomothetic models.*** *Box plots of MAPE distribution of feature-based model* ***(A)*** *and log2 fold change in CAT-DI prediction accuracy of feature-based model over the baseline model* ***(B)*** *across individuals for different latent depression traits (panel), modeling approaches (color), CAT-DI regression model (x-axis), and feature imputation methods (transparency). All plots are based on individuals with at least five assessments in the test set (N=143). The dark black line represents the median value; the box limits show the interquartile range (IQR) from the first (Q1) to third (Q3) quartiles; the whiskers extend to the furthest data point within Q1-1.5*IQR (bottom) and Q3+1.5*IQR (top). LOCF: last observation carried forward. CS(xdf): cubic spline with x degrees of freedom. CS(cv): best-fitting cubic spline according to leave-one-out cross-validation.*

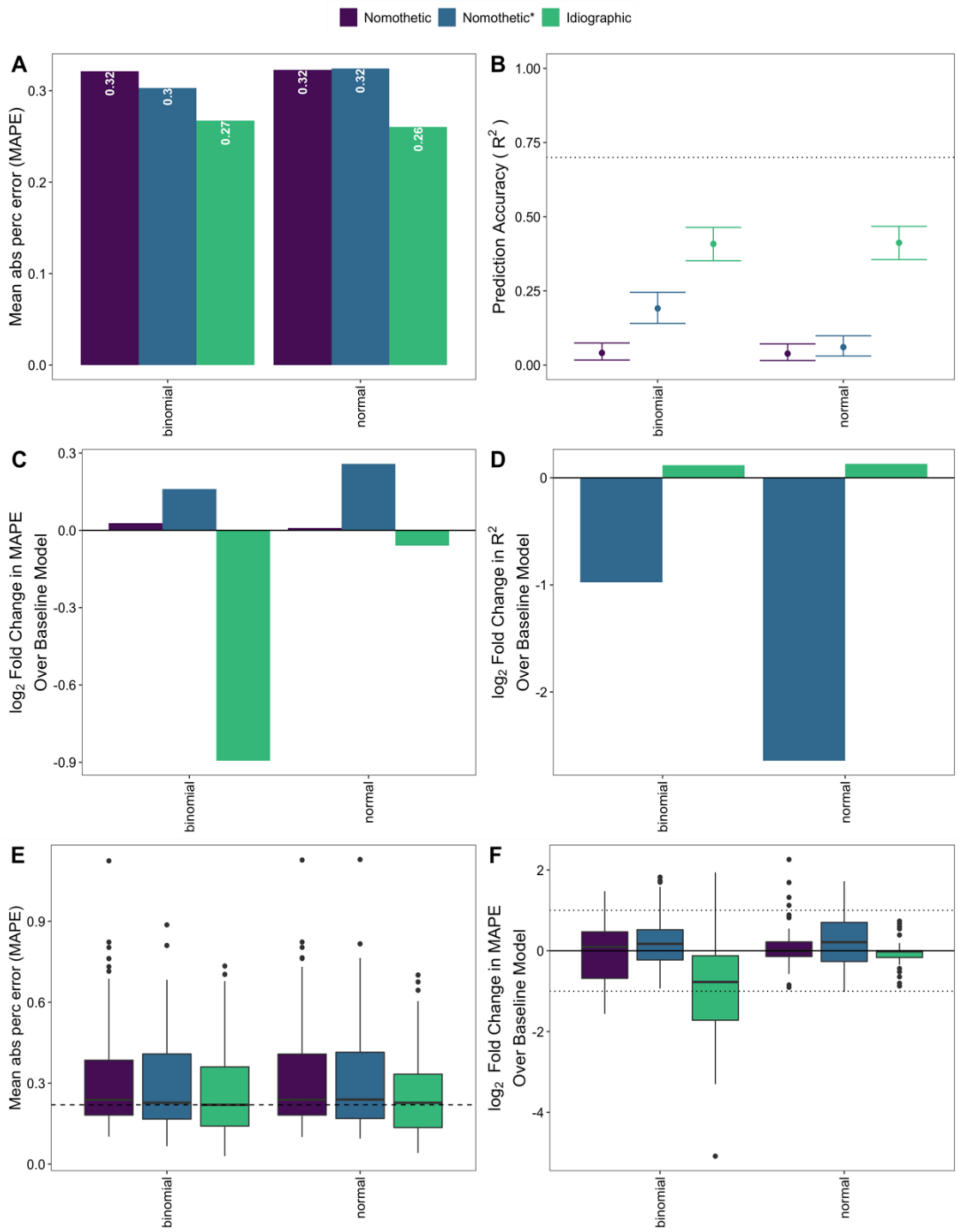


*Sup Figure 8:* ***Idiographic models achieve higher group and individual level prediction accuracy than nomothetic models when CAT-DI is modeled at the (bi)weekly level..(A-D)*** *Population-level CAT-DI prediction accuracy of the feature-based model (A-B) and log2 fold change in population-level CAT-DI prediction accuracy of the feature-based model over the baseline model (C-D) as measured by MAPE (A and C) and* $R^2$ *(B and D) across all individuals in the test set for different modeling approaches (color) and CAT-DI regression model (x-axis). The dotted line in B indicates 70% prediction accuracy and the bars indicate 95% confidence intervals of* $R^2$. ***(E-F)*** *Box plots of MAPE distribution of feature-based model* ***(E)*** *and log2 fold change in CAT-DI prediction accuracy of feature-based model over the baseline model* ***(F)*** *in the test set across individuals for different modeling approaches (color) and CAT-DI regression model (x-axis). The dark black line represents the median value; the box limits show the interquartile range (IQR) from the first (Q1) to third (Q3) quartiles; the whiskers extend to the furthest data point within Q1-1.5*IQR (bottom) and Q3+1.5*IQR (top). Plots E-F are based on individuals with at least five assessments in the test set (N=143). For all six plots, features were imputed using AutoComplete. MAPE: mean absolute percentage error.*
